# Novel protein-altering variants associated with serum apolipoprotein and lipid levels

**DOI:** 10.1101/2021.09.19.21263610

**Authors:** Niina Sandholm, Ronja Hotakainen, Jani K Haukka, Fanny Jansson Sigfrids, Emma H Dahlström, Anni Antikainen, Erkka Valo, Anna Syreeni, Elina Kilpeläinen, Anastasia Kytölä, Aarno Palotie, Valma Harjutsalo, Carol Forsblom, Per-Henrik Groop, on behalf of the FinnDiane Study Group

## Abstract

Dyslipidemia is a major risk factor for cardiovascular disease. While common genetic variants are known to modestly affect the serum lipid concentrations, rare genetic mutations can cause monogenic forms of hypercholesteremia and other genetic disorders of lipid metabolism. Aiming to identify low-frequency protein-altering variants (PAVs) affecting lipoprotein and lipid traits, we analyzed whole-exome and whole-genome sequencing data of 481 and 573 individuals with type 1 diabetes, respectively. The phenotypic data consisted of 97 serum lipid, apolipoprotein, or other metabolic phenotypes obtained with clinical laboratory measurements and nuclear magnetic resonance (NMR) technology. Single variant analysis identified a novel association between *LIPC* p.Thr405Met (rs113298164) and serum apolipoprotein-A1 levels (p=7.8×10^−8^). In the *APOB* gene, we identified novel associations at two protein-truncating variants (PTVs) resulting in lower serum apolipoprotein B levels (p=5.6×10^−4^). The burden of PAVs was significantly associated with lipid phenotypes in *LIPC, RBM47, TRMT5*, and *GTF3C5* (p<2.9×10^−6^). The *RBM47* gene is required for apolipoprotein-B post-translational modifications, and in our data, the association between *RBM47* and apolipoprotein C-III levels was led by a rare 21 base pair Ala496-Ala502 deletion; as replication, the burden of rare deleterious variants in *RBM47* was associated with TG-to-HDLC ratio in WES of 20,917 individuals (p=0.0093). Two PAVs in *GTF3C5* were highly Finnish-enriched and associated with cardiovascular phenotypes in external data, whereby the *TRMT5* p.Ser185Cys lead variant was associated with stroke phenotypes. Altogether, we identified both novel variant associations in known lipid genes, as well as novel genes implicated in lipoprotein metabolism.

## Introduction

Cardiovascular disease (CVD) is the leading cause of mortality worldwide.^1^ Plasma lipid concentrations are key CVD risk factors, and thus, lipid-lowering medication is an essential treatment option to prevent CVD. Genetic factors explain approximately 10-54% of plasma lipid levels^2^ and the largest genome-wide association study (GWAS) on plasma lipid values identified nearly 400 genetic loci associated with plasma low-density lipoprotein cholesterol (LDLC), triglycerides (TG), total cholesterol, or high-density lipoprotein cholesterol (HDLC).^3^ Genome-wide analyses focusing on the exonic regions of the genome have identified low-frequency or rare PAVs that contribute to the previously observed common variant associations, or even explain most of the associations observed for those.^4, 5^ The low-frequency protein-altering variants (PAVs) can have a much stronger impact on the phenotype than the disease-associated common genetic variants, which are enriched for gene regulatory variants and often have moderate effect sizes.^6^ Furthermore, identification of rare loss-of-function variants may reveal genes that can be targeted to prevent a disease, such as the LDLC-lowering loss-of-function variants in *PCSK9* (MIM:607786), the identification of which resulted in the PCSK9 inhibitors for preventing CVD.^7^

Lipidomic profiles consisting of more detailed lipid and lipoprotein subtypes can increase our understanding of the complex lipidomic regulatory networks and, occasionally, outperform the traditional lipid variables in risk prediction.^6^ In addition, apolipoprotein concentrations provide added awareness of the burden of circulating lipoproteins. For example, one apolipoprotein-B (apoB) molecule is embedded in each very-low-density lipoprotein (VLDL), intermediate-density lipoprotein (IDL), low-density lipoprotein (LDL), and lipoprotein(a) (Lp[a]) particle, and apoB seems to estimate the atherogenic risk more accurately than the traditional LDLC^8^ or even multivariable data-driven sub-grouping of lipoprotein subtypes.^9^ Furthermore, apolipoprotein C-III (apoC-III) – found particularly in the triglyceride-rich lipoproteins (TRLs) – has been recently implicated as a CVD risk factor both in the general population and in individuals with type 1 diabetes.^10, 11^ Genetic studies of these refined lipid phenotypes have revealed common variants contributing e.g. to apoB levels^12^, but also identified rare genetic factors with drastic impact e.g. on apoC-III levels, reflected on the CVD risk.^13^

Individuals with type 1 diabetes carry a considerable CVD risk burden, with a 7.5-fold incidence ratio for coronary artery disease (CAD) *vs*. the general population; in the presence of diabetic kidney disease (DKD), this ratio is up to 27-fold.^14^ Dyslipidemia – particularly hypertriglyceridemia – is an established risk factor for both CAD and DKD in these individuals.^15^ The incidence of CAD increases already below the currently recommended TG cut-off of 1.7 mmol/L, suggesting that the additional risk imposed by lipids is pronounced in diabetes. Therefore, genetic studies on lipids in type 1 diabetes are of particular importance. Furthermore, the Finnish population provides advantages and increased statistical power for studying rare variants, as some deleterious rare variants are present at higher frequencies in Finnish subjects due to population isolation and recent genetic bottlenecks.^16^ Therefore, using whole-exome and whole-genome sequencing (WES and WGS, respectively), we aim to identify novel PAVs and protein-truncating variants (PTVs, as putative loss-of-function variants) affecting serum lipid and lipoprotein measurements, complemented with serum nuclear magnetic resonance (NMR) measurements in Finnish individuals with type 1 diabetes.

## Material & Methods

### Patients

The Finnish Diabetic Nephropathy Study (FinnDiane) is an ongoing nationwide prospective study, established in 1997 to pinpoint risk factors for long-term diabetic complications. The study currently comprises more than 5,200 individuals with type 1 diabetes. The study protocol was approved by the Ethical Committee of the Helsinki and Uusimaa Hospital District and local ethics committees and participants gave their informed consent before recruitment. This study was performed following the Declaration of Helsinki.

### Phenotypes

Type 1 diabetes was defined as an onset of diabetes before the age of 40 and the initiation of permanent insulin treatment during the first year after diagnosis. The clinical characterization of the participants and the recruitment has been described earlier.^17^ In brief, data on diabetic complications, history of cardiovascular event(s), and prescribed medications were registered using standardized questionnaires, and blood and urine samples were collected during a standard visit to the attending physician. DNA was extracted from blood.

Blood serum lipid and apolipoprotein concentrations were determined at the central research laboratory (CL) of Helsinki University Central Hospital, Finland,^15^ with detailed methods in **Supplementary Table S1**.

Proton NMR spectroscopy was utilized to quantify numerous lipoprotein subclasses and their contents along with several metabolites from the blood serum of 3,544 participants from the FinnDiane Study. Lipoproteins were classified according to their diameter into VLDL, IDL, LDL, and HDL particles. These were further subdivided as described earlier.^18^ The spectroscopy was tailored to target three molecular windows: the lipoprotein lipids, low molecular weight compounds,^19^ and serum lipid extracts.^20^ The NMR spectroscopy was performed in four different batches, and some metabolites, e.g., amino acids, were determined only in one batch.

Lipid-lowering medication use, defined as statin use, was accounted for by using a similar approach previously adopted by others.^4, 21^ We divided total cholesterol by 0.8 to account for the 20% reduction in serum total cholesterol induced by statins.^22^ We used this adjusted value to calculate LDLC with the Friedewald formula.^23^ We divided the subgroups of NMR-measured LDLC by 0.7 to account for a 30% reduction in LDLC. As statins also affect VLDL particles, the NMR VLDL cholesterol measurements were divided by 0.8.^24^ We left serum TG and HDL cholesterol (HDLC) measurements unadjusted, as heritability estimates do not significantly improve when adjusting for statin use.^25^ All other lipid variables were left unadjusted, as the exact effect of statins remains unclear.

The kidney status was based on albuminuria status, and subjects were classified as having normal albumin excretion rate (AER <20 μg/min), microalbuminuria (20–199 μg/min), macroalbuminuria (≥200 μg/min), or renal failure requiring dialysis or kidney transplant. CAD was based on Statistics Finland and the National Care Register for Health Care using the ICD-10 codes I21, I22, and I23 for myocardial infarction; and the Nordic Classification of Surgical Procedure codes for coronary bypass surgery or coronary balloon angioplasty.

### Whole exome- and whole-genome sequencing data

WES data were available for 481 individuals with either normal AER despite long (≥32 years) diabetes duration, or with severe DKD, i.e. macroalbuminuria and/or renal failure. The sequencing process, variant calling, annotation, and quality control have been described earlier.^26, 27^ In brief, sequencing was performed with Illumina HiSeq2000 platform at the University of Oxford, UK, with an average requirement of 20x target capture with an above 80% coverage, resulting in mean sequencing depth of 54.97 bases per position. Variant calling was performed with Genome analysis toolkit (GATK) v2.1, with human genome assembly GRCh37 as a reference. Variants were updated to the GRCh38 assembly using the UCSC liftOver tool with default parameters and a hg19 to hg38 chain file.

WGS data were available for 598 unrelated participants from the FinnDiane Study, non-overlapping with the WES individuals. Similar to WES, the WGS data included 292 controls with normal AER and long diabetes duration (≥35 years) and 291 cases with severe DKD. The sequencing was performed using an Illumina HiSeq X platform (Macrogen Inc., Rockville, MD, USA). Variant calling was done using Broad Institute’s best practices guidelines with GATK v4.^28^ The humangenome assembly GRCh38 was used as a reference. Variants were filtered to those with variant call rate >98% and in Hardy Weinberg equilibrium (HWE; *p*-value >10^−10^, or >10^−50^ in HLA region, as all had type 1 diabetes). The final data included 21.92 million variants. A total of 573 samples passed the quality control filters, including the percentage of mapped de-duplicated reads and excess heterozygosity. Principal component analysis indicated no population outliers. Lipid-related phenotypes were available for 474 individuals.

All WGS and WES variants were annotated for their functional effects with SnpEff v4.3^29^ and GrCh38.86 database. Variants classified by SnpEff as PTV (exon loss, frameshift, stop or start gained or lost, splice acceptor, and donor variants) and PAVs (PTV plus missense variants, and inframe insertions or deletions) were included in the analyses.

### Single variant analysis for WES and WGS variants

All PAVs were tested for association with the lipid, apolipoprotein, or other metabolic phenotypes, separately for WES and WGS data sets, using the Rvtests score test.^30^ Analyses were adjusted for sex, age, and the two first genetic principal components. The NMR measured phenotypes were additionally adjusted for the NMR measurement batch. Inverse normal transformation was performed for all trait residuals. Finally, single-variant meta-analysis of WES and WGS cohorts was performed with RAREMETAL^31^ (**Figure 1**). Exome-wide significance was defined as <4.3×10^−7^, adjusted for 116,567 tested variants (Bonferroni correction for multiple testing with α=0.05 significance level). P-values < 1×10^−5^ were considered suggestive. Detailed single-variant statistical analyses, including survival models for CVD phenotypes, were performed in R using the survival package.^32^

**Figure 1:**
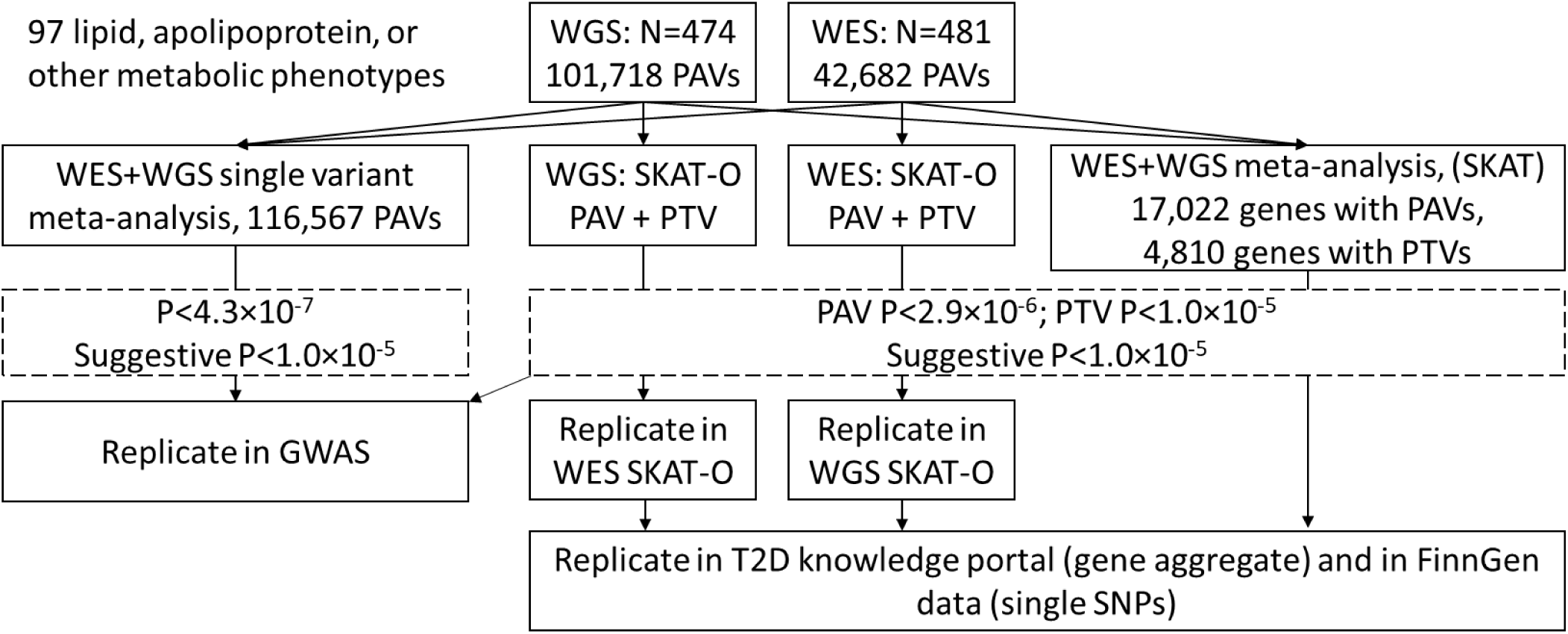
Flow-chart of the study design. PTVs: protein truncating variants i.e. exon loss, frameshift, stop or start gained or lost, splice acceptor, and donor variants. PAVs: protein altering variants, defined as PTV plus missense variants, inframe insertions and deletions.

### Single variant replication

Variants with a P-value <1×10^−5^ from the single variant meta-analysis were chosen for replication in FinnDiane GWAS data with 6,449 individuals, genotyped with Illumina HumanCoreExome Bead arrays, genotypes called with zCall algorithm, and initial quality control performed at the University of Virginia.^33^ Genotyping data were lifted over to build version 38 (GRCh38/hg38), and data from the four genotyping batches were merged. In sample-wise quality control, individuals with high genotype missingness (>5%), excess heterozygosity (±4 standard deviations), and non-Finnish ancestry (none) were removed. In variant-wise quality control variants with high missingness (>2%), low HWE p-value (<10^−6^), or minor allele count [MAC] <3 were removed. Chip genotyped samples were pre-phased with Eagle 2.3.5, and genotype imputation was performed with Beagle 4.1 (version 08Jun17.d8b) based on the population-specific SISu v3 imputation reference panel with WGS data for 3,775 Finnish individuals; only variants with high imputation quality of r^2^>0.8 were included. Depending on the phenotype, data were available for 820 (tyrosine) to 4,653 (total cholesterol) individuals after excluding the FinnDiane WES and WGS individuals to ensure independent replication. Rvtests software was used, and analyses were adjusted for sex, age, and the kinship matrix. The variants were further tested for association with any DKD (micro- or macroalbuminuria or renal failure vs. normal AER), severe DKD (macroalbuminuria or renal failure vs. normal AER), and renal failure vs normal AER. For CAD, we used pre-calculated GWAS results from FinnDiane.^34^

We used Sanger sequencing to confirm the 21bp deletion in the *RBM47* gene in seven heterozygotes with lipid data. We designed the primers with Primer3 software^35^ and ordered them from Sigma-Aldrich Company Ltd (Haverhill, UK), and sequencing was performed at FIMM (Institute for Molecular Medicine Finland, Helsinki, Finland).

### WES and WGS gene-based analysis

Gene-based tests were performed for WES and WGS data using optimized sequence kernel association test (SKAT-O).^36^ We analyzed the burden of PAVs or PTVs with MAF < 5% using Rvtests (v. 2019-02-09) --kernel skato option. Analyses were adjusted for age, sex, and genetic principal components. NMR phenotypes were further adjusted for the measurement batch. Statistical significance for the burden of PAVs and PTVs were defined as 2.9×10^−6^ and 1.0×10^−5^, respectively (adjusted for up to 17,022 genes with PAVs, and 4,810 genes with PTVs in the WES-WGS meta-analysis; Bonferroni correction with α=0.05). Significant WES SKAT-O results were internally replicated with WGS SKAT-O results, and *vice versa* (**Figure 1**). Significant replication was defined as P<0.05.

Meta-analysis of the gene-based enrichment of PAVs and PTVs in WES and WGS data was performed with SKAT^37^ implemented in RAREMETAL,^31^ and based on the single-variant score test results (described above) and covariance matrices from Rvtests.^30^ The pooled variants were re-annotated with the anno tool in RAREMETAL before analysis. Again, variants were limited to those with MAF <5% and analyzed for all PAVs, or PTV variants only. Significant burden of PAVs or PTVs were defined as for WES and WGS SKAT-O analysis (**Figure 1**).

### Replication of gene aggregate findings

Replication for gene aggregate findings was sought from the AMP type 2 diabetes (T2D)-GENES exome sequencing study on various metabolic traits including the traditional lipid variables.^38^ Variant associations with cardiovascular endpoints for the single variants within the gene aggregate findings were sought from the FinnGen study GWAS results based on 109 “Diseases of the circulatory system” phenotypes constructed from ICD codes for 176,899 Finnish individuals (freeze 4, accessed 11 March 2021), imputed with a Finnish reference panel with 3,775 individuals. Variant enrichment estimates in the Finnish population vs. the gnomAD non-Finnish-non-Estonian European samples were available in the same data.

### Functional annotation

Ensembl Variant Effect Predictor was used to predict the effect of the identified variants, based on SIFT and PolyPhen scoring. Gene expression in various tissues was used to annotate identified genes and studied in Human Protein Atlas.

## Results

The WES and WGS data included 42,682 and 101,718 PAVs, respectively, available for participants with lipid data (**Supplementary Table S2**); 79-82% were low-frequency variants with MAF<5%. A total of 2,240 and 9,577 variants in WES and WGS, respectively, were annotated as PTV likely to disrupt the protein structure; defined here as frameshift, stop or start gained or lost, exon loss, or splice site acceptor and donor variants. The vast majority, 82-90% of the PTVs, had MAF<5%.

### Single variant association analysis

In the WES-WGS meta-analysis, a missense variant rs113298164 (Thr405Met, MAF 1.7%) in *LIPC* (MIM:151670) was associated with higher serum apoA-1 levels (p=7.8×10^−8^; **Table 1, Figure 2A**). In Thr405Met carriers (n=31), the median serum apoA-1 was 163 mg/dl (inter-quartile range [IQR] 145 - 183) mg/dl, vs. 138 (IQR 121 - 153) mg/dl in the non-carriers (multivariate ANOVA p=1.46×10^−9^). However, in Cox proportional-hazard models, Thr405Met was not associated with CAD, nor with stroke (**Figures 2B, 2C**).

**Table 1:**
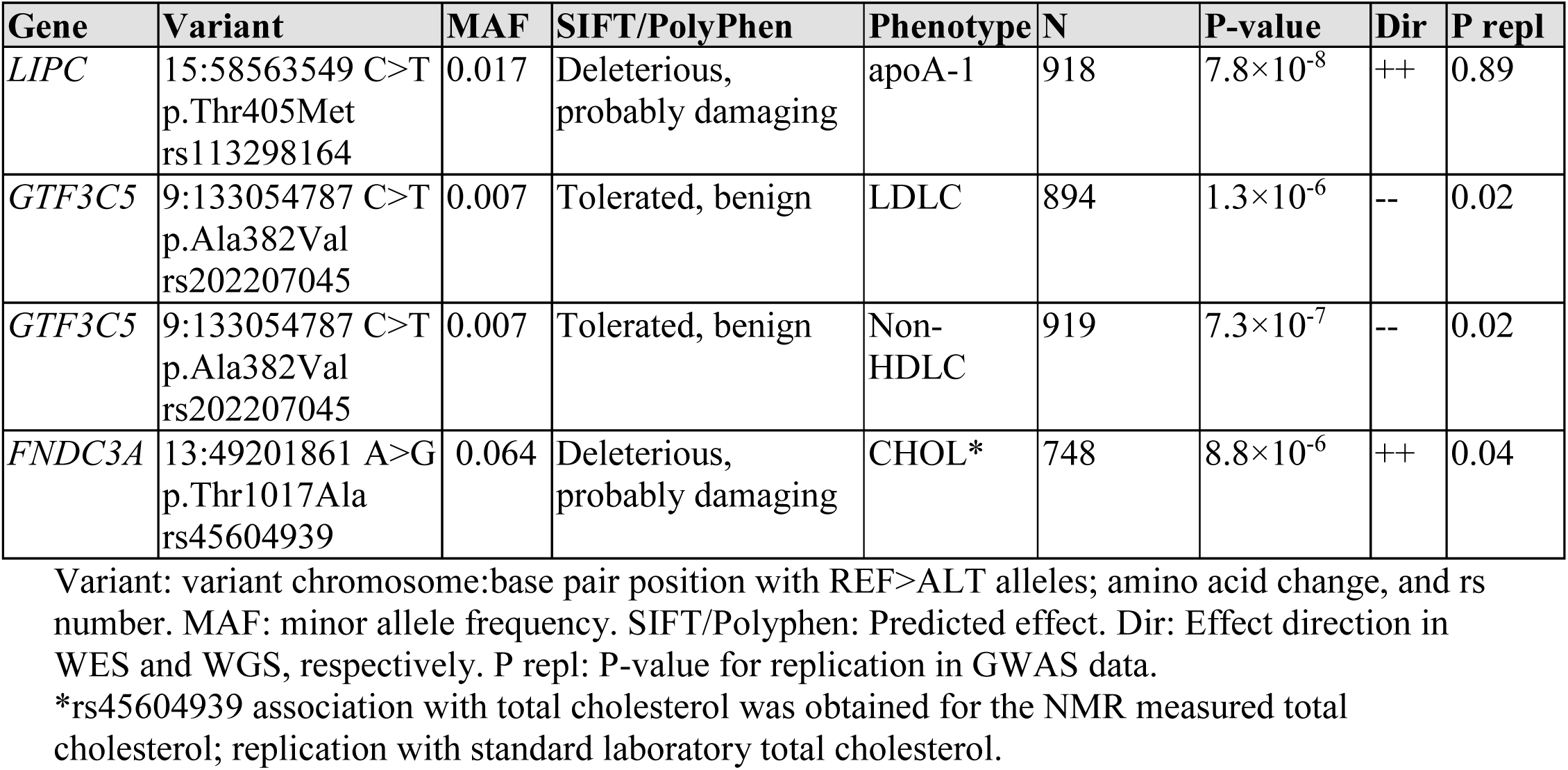
Single variant association results for variants reaching exome-wide significance (p<4.3×10_-7_) or with evidence of replication (p<0.05).

**Figure 2:**
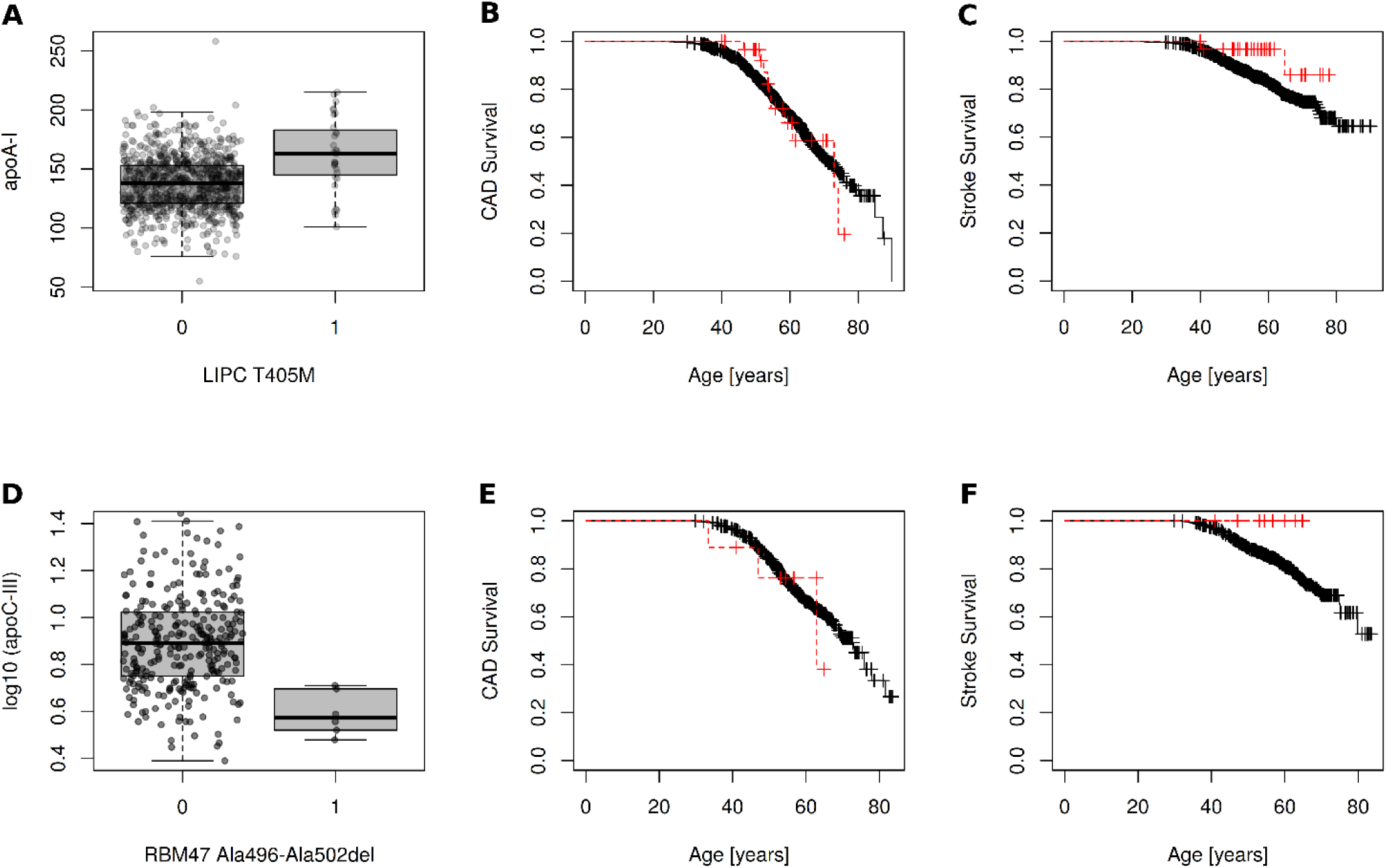
Rare variants in *LIPC* and *RBM47* are associated with serum apolipoprotein A1 and C3 levels, respectively, but not with cardiovascular events. For *LIPC* Thr405Met (rs113298164) data is combined for WES and WGS; *RBM47* Ala496-Ala502del is only found in the WGS data. **A:** *LIPC* Thr405Met (rs113298164) is associated with higher apoA-1 (p=7.8×10^−8^; multivariate ANOVA p= 1.46×10^−9^; N=887 carriers, 31 non-carriers). Group number in **A** and **D** indicates the number of rare variants, i.e. 0 refers to non-carriers, 1 refers to heterozygous variant carriers. **B and C:** Kaplan-Meier survival plot of CAD events (**B**) and strokes (**C**), stratified by *LIPC* Thr405Met variant count: black lines indicate non-carriers, red dashed line indicates variant carriers. Cox proportional hazard (PH) model *p*-value was non-significant for both CAD and strokes. **D:** Serum apoC-III levels are reduced in the *RBM47* Ala496-Ala502del carriers (p=2.49×10^−6^, multivariate ANOVA p= 2.92×10^−4^; N = 288 non-carriers, 6 carriers). **E and F:** Kaplan-Meier survival plot of CAD events (E) and strokes (F), stratified the *RBM47* Ala496-Ala502del variant number (Cox PH p-value non-significant).

Furthermore, 26 variants were suggestively associated with lipid, apolipoprotein, lipoprotein and, metabolite phenotypes (p<1×10^−5^; **Supplementary Table S3**). One of the variants was a 21bp inframe deletion in *RBM47* (MIM:619104; Ala496-Ala502del, rs564837143, MAF=1.0%, p=2.5×10^−6^) found in the WGS data only, and associated with lower serum apoC-III concentrations, with median apoC-III of 3.74 (IQR=3.38-4.69) mg/dl in the six Ala496-Ala502del carriers vs. 7.79 (IQR=5.62-10.51) mg/dl in the non-carriers (**Figure 2D**). In the subsequent analysis of the full WGS data with nine Ala496-Ala502del carriers (with or without apoC-III available), three experienced a CAD event during the full study period, not significantly different from the non-carriers (**Figure 2E-F**).

### Replication of putative associations

The FinnDiane GWAS dataset contained 26 of the 27 lead variants with high imputation quality.^39^ Two of these were replicated with nominal significance: p.Thr1017Ala (rs45604939) in *FNDC3A* (MIM:615794) was associated with higher total cholesterol (MAF=0.063, p=0.04); and p.Ala382Val (rs202207045) in *GTF3C5* (MIM:604890) with lower LDLC and non-HDLC (MAF 0.008, p=0.02 for both; **Table 1, Supplementary Table S3**). In the independent FinnGen GWAS data of 106 circulatory phenotypes for 176,899 Finnish individuals, rs45604939 in *FNDC3A* was nominally associated (p<0.05) with multiple circulatory phenotypes.

### WES and WGS gene-based analysis

We performed gene aggregate tests to identify genes enriched for low frequency (MAF≤5%) PAVs and PTVs. In WES, PAVs in *AKAP3* (MIM:604689) were significantly associated (p<2.9×10^−6^, adjusted for 17,022 genes) with the TG content of the extremely large VLDL particles (p=1.4×10^−7^; **Table 2**). Furthermore, PTVs in *PTGER3* (MIM:176806) were significantly associated (p<1.0×10^−5^, adjusted for 4,810 genes) with free cholesterol in medium-sized HDL particles (p=9.8×10^−6^). Two additional genes reached a suggestive p-value <1×10^−5^ for PAVs (**Table 2**). In WGS, SKAT-O analysis revealed that PAVs in *RBM47* were associated with serum apoC-III concentrations (p=2.2×10^−6^). Of note, the association was driven by the 21 bp inframe deletion of the *RBM47* gene identified in the WGS single variant analysis. Furthermore, in WGS, PTVs in *SBDS* (MIM:607444) were also associated with serum apoC-III concentrations (stop gain, and a splice donor variant; p=5.0×10^−6^). Finally, a splice donor PTV in the *DEFT1P/DEFT1P2* genes was associated with phospholipids in extra-large VLDL particles (p=1.3×10^−6^). Four additional genes had PAVs suggestively associated with lipid phenotypes (p<1×10^−5^; **Table 2**).

**Table 2:** WES and WGS SKAT-O gene burden test results for genes reaching a suggestive p-value <1×10_-5_, and the internal replication in the other data set. In the exome-wide discovery, p-values <2.9×10^−6^ for PAVs, and p-values <10^−5^ for PTV burden was defined as statistically significant, highlighted in italics; in replication, p-values <0.05 are highlighted in italics

| Gene | Gene position | Type | Pheno | Discovery |  |  | Replication |  |  |
| --- | --- | --- | --- | --- | --- | --- | --- | --- | --- |
|  |  |  |  | N | Nvar | P | N | Nvar | P |
| Discovery study: WGS |  |  |  |  |  |  |  |  |  |
| DEFT1P/ |  |  |  |  |  |  |  |  |  |
| DEFT1P2 | 8:7006280-7008824 | PTV | VLDLPL XL | 187 | 1 | $1.3\times10^{-6}$ | | | |
| SBDS | 7:66987676-66995696 | PTV | apoC-III | 294 | 2 | $5.0\times10^{-6}$ | | | |
| RBM47 | 4:40423254-40629866 | PAV | apoC-III | 294 | 3 | $2.0\times10^{-6}$ | 323 | 1 | 0.398 |
| TRMT5 | 14:60971448-60981690 | PAV | CHOL | 329 | 8 | $3.5\times10^{-6}$ | 419 | 4 | 0.322 |
| TRMT5 | 14:60971448-60981690 | PAV | VLDLPL XS | 329 | 8 | $5.9\times10^{-6}$ | 419 | 4 | **0.015** |
| TRMT5 | 14:60971448-60981690 | PAV | IDLFC | 329 | 8 | $6.8\times10^{-6}$ | 419 | 4 | **0.019** |
| TRMT5 | 14:60971448-60981690 | PAV | LDLC L | 329 | 8 | $9.3\times10^{-6}$ | 419 | 4 | 0.436 |
| CCAR1 | 10:68721143-68792377 | PAV | VLDL XL | 329 | 2 | $7.4\times10^{-6}$ | 419 | 3 | 0.555 |
| CYP3A43 | 7:99828012-99866106 | PAV | LDLCE M | 329 | 6 | $8.7\times10^{-6}$ | 419 | 3 | 0.067 |
| CYP3A43 | 7:99828012-99866106 | PAV | LDLCE L | 329 | 6 | $8.7\times10^{-6}$ | 419 | 3 | **0.038** |
| CYP3A43 | 7:99828012-99866106 | PAV | LDL M | 329 | 6 | $9.2\times10^{-6}$ | 419 | 3 | 0.074 |
| DIPK1A | 1:92832728-92961522 | PAV | TG | 451 | 4 | $9.9\times10^{-6}$ | 469 | 3 | 0.848 |
| Discovery study: WES |  |  |  |  |  |  |  |  |  |
| PTGER3 | 1:70852352-71047808 | PTV | HDLFC M | 419 | 1 | $9.8\times10^{-6}$ | 329 | 1 | 0.119 |
| AKAP3 | 12:4615507-4649047 | PAV | VLDLTG XXL | 304 | 5 | $6.0\times10^{-7}$ | 245 | 5 | 0.766 |
| TTYH1 | 19:54415430-54436719 | PAV | HDLCE L | 419 | 3 | $5.7\times10^{-6}$ | | | |
| ATP4A | 19:35550192-35563658 | PAV | HDLTG XL | 419 | 3 | $8.1\times10^{-6}$ | 329 | 3 | 0.810 |

Discovery: Exome-wide analysis of either WGS, or WES data. Replication: for WGS, replication in WES data; for WES, replication in WGS data. Gene position: Chromosome number:start-end; If multiple gene isoforms exist, only one set of co-ordinates are given. Type: burden of PAVs or PTVs. Nvar: number of variants of the given variant type in the gene in discovery/ replication data. apoC-III: Serum Apolipoprotein C-III, CHOL: Total cholesterol, HDLCE L: Cholesterol ester in large HDL, HDLFC M: Free cholesterol in medium HDL, HDLTG XL: TG in extra large HDL, IDLFC: Free cholesterol in IDL particles, LDL M: Total lipids in medium LDL, LDLCE L: Cholesterol in large LDL, LDLCE M/L: Cholesterol esters in medium/large LDL, VLDL XL: Total lipids in extra large VLDL, VLDLPL XS/XL: Phospholipid in extra small/ extra large VLDL, VLDLTG XXL: TG in extremely large VLDL.

We sought replication of the suggestive SKAT-O results by performing an internal replication between the two data sets. The PAVs of the *TRMT5* (MIM: 611023) gene were associated in WGS with free cholesterol in IDL particles (p=6.8×10^−6^) and with phospholipids in extra small VLDL particles (p=5.9×10^−6^), and these associations were replicated in WES (p=0.019 and p=0.015, respectively; **Table 2**). In addition, the association between PAVs in *CYP3A43* (MIM:606534), and cholesterol esters in large LDL particles in WGS (p=8.7×10^−6^), was replicated in WES (p=0.038).

Finally, to increase the statistical power, we performed gene aggregate analysis in the combined WES and WGS data by applying SKAT meta-analysis implemented in RAREMETAL. The burden of PAVs was significantly associated (p<2.9×10^−6^) with lipid phenotypes in four genes (*LIPC, RBM47, TRMT5*, and *GTF3C5* **Table 3**): PAVs in the *LIPC* gene – including rs113298164 from the single variant meta-analysis – were associated with serum apoA-1 levels (p=1.48×10^−7^). The PAVs in *RBM47* were associated with serum apoC-III concentrations also in the WES-WGS SKAT meta-analysis (p=1.33×10^−6^), and PAVs in *TRMT5* were associated with phospholipids in extra small VLDL particles (p=7.87×10^−7^). The *TRMT5* PAVs were nominally associated also with multiple IDL phenotypes (**Figure 3**). Finally, PAVs found in the *GTF3C5* gene were associated with total cholesterol, LDLC, and non-HDLC. SKAT meta-analysis on DKD and CVD for the lead genes suggested that PAVs in *CYP3A43* are associated with DKD (p=0.004, rank 43/17,578 genes i.e. top 0.3%: **Supplementary Table S4**).

**Table 3:**
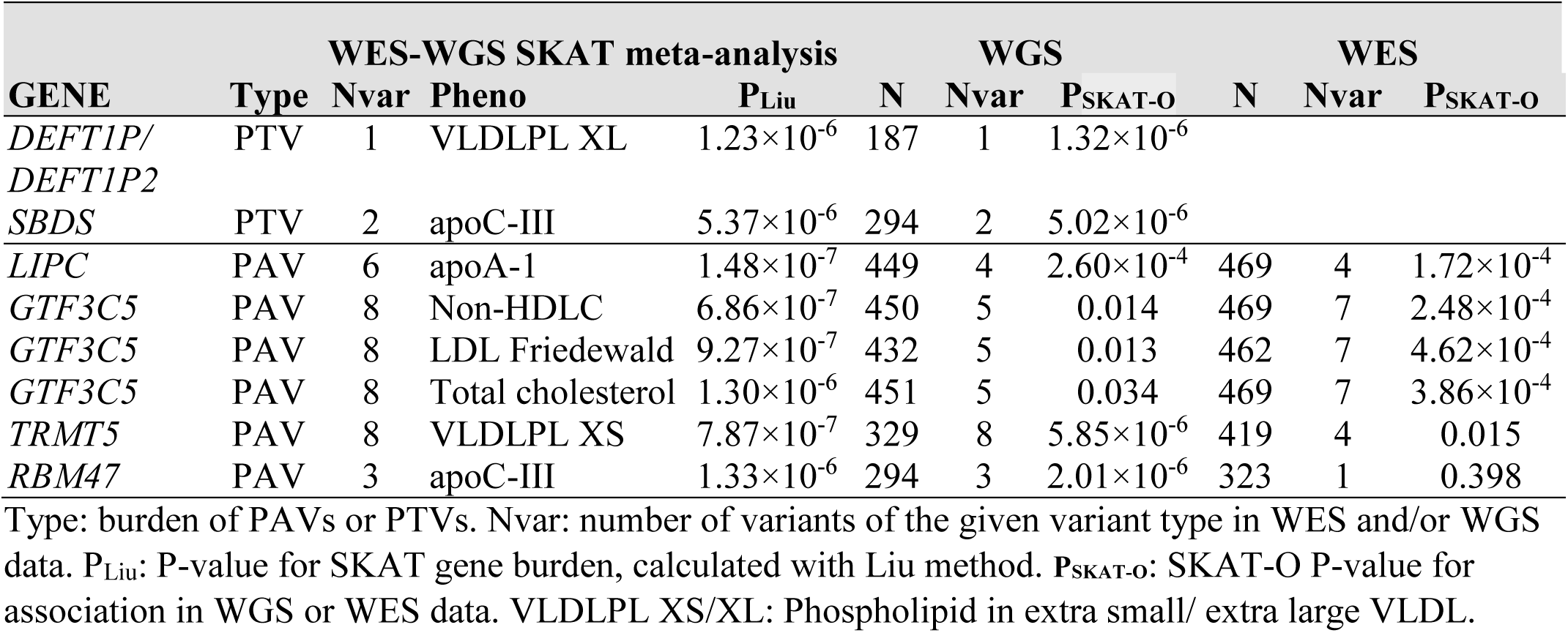
WES and WGS SKAT meta-analysis results for genes with significant burden of PAVs (p-value < 2.9×10_-6_) or PTVs (p-value < 1×10_-5_). Also indicated the P-values from the separate WGS and WES SKAT-O gene burden analyses.

**Figure 3:**
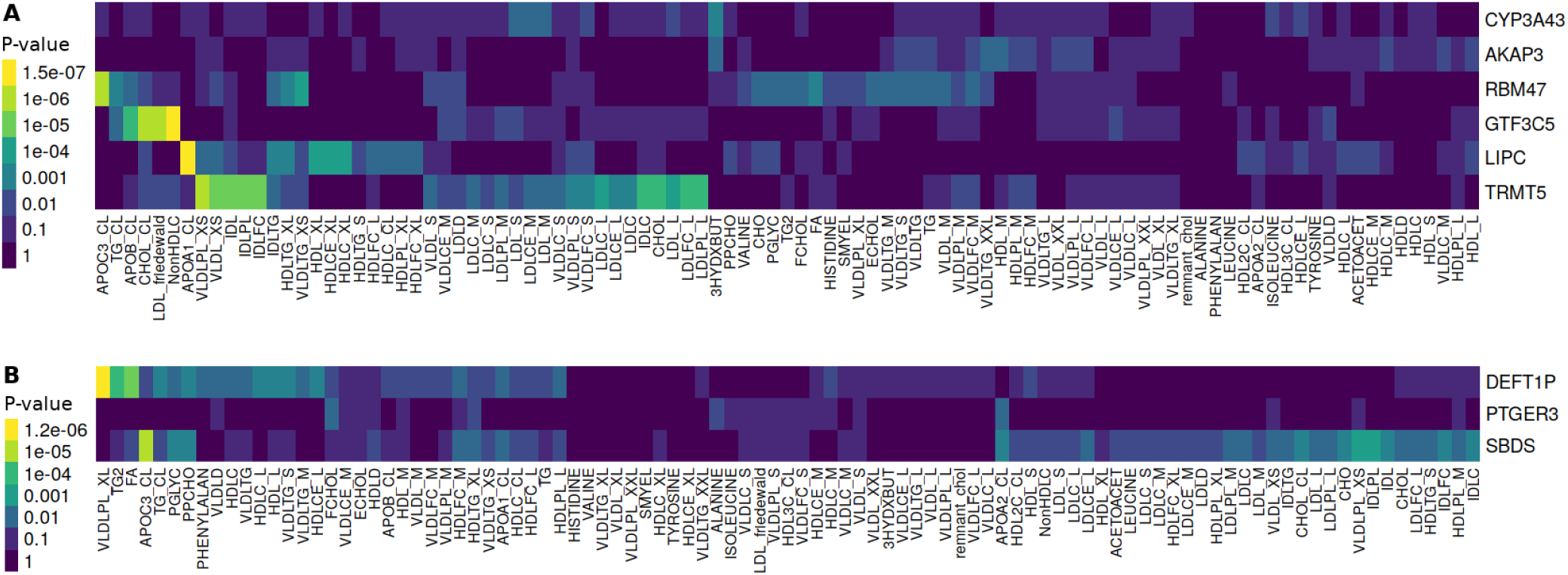
WES-WGS SKAT meta-analysis results across all the studied phenotypes for the lead genes. The color indicates the strength of the gene – phenotype association (SKAT *p*-value). **A)** WES-WGS PAV SKAT meta-analysis across phenotypes. B) WES-WGS PTV SKAT meta-analysis across phenotypes. The phenotypes are ordered according to their similarity in clustering of the association data.

Among the 33 PAVs found in these lead genes, 20 were found with good imputation quality in the FinnDiane GWAS data. In addition to the above-mentioned *GTF3C5* rs202207045 variant association (p=0.02 for LDLC and non-HDLC), a *LIPC* p.Phe368Leu (rs3829462) variant was associated with higher apoA-1, along with a rare (MAF=0.0002, MAC=1.5) low imputation quality (0.37) *LIPC* p.Ser301Phe variant (p=0.04; **Supplementary Table S5**).

Validation of the gene aggregate results was sought from the AMP T2D-GENES exome sequence gene-level analysis of rare variants.^40^ Variants in *LIPC* were associated with HDLC (p=2.72×10^−14^) and other lipid traits (p<0.05); variants in *RBM47* with TG-to-HDLC ratio (p=0.0093 for variants predicted deleterious by 5 methods [“5/5”]); and variants in *PTGER3* with total cholesterol (5/5 p=0.0033) and LDLC (5/5 p=0.0045).

We further sought replication for the individual variants contributing to the gene aggregate associations from the GWAS data of the FinnGen study. The strongest evidence of replication was found for a rare (MAF=0.004), 80-fold Finnish-enriched deleterious start-lost variant rs189383196 in *GTF3C5*, associated with non-ischemic cardiomyopathy (p=2.8×10^−5^), hypertension (p=6.7×10^−4^), and 18 other circulatory phenotypes (p<0.05; **Supplementary Table S6**). Also, another rare (MAF=0.001), 77-fold Finnish-enriched, deleterious rs369889499 (p.Tyr347Cys) variant in *GTF3C5* was associated with multiple phenotypes, including angina pectoris (p=9.20×10^−5^) and ischemic heart disease (p=6.10×10^−4^). In the *TRMT5* gene, the variant with the strongest individual association, rs115400838 (p.Ser185Cys), was associated with multiple stroke phenotypes, e.g. “Stroke, excluding subarachnoid hemorrhage”, p=1.90×10^−4^.

### Association for genes causing monogenic forms of dyslipidemia

Previously, rare variants in multiple genes have been associated with severe monogenic forms of dyslipidemia. We studied the PAV and PTV burden in eight genes causing hypercholesteremia, five genes causing monogenic hypertriglyceridemia, and seven genes causing genetic disorders of HDL metabolism, including the *LIPC* gene (**Supplementary Table S7**).^41^ In the hypercholesteremia-causing *APOB* (MIM:107730), we identified two frameshift PTVs in exon 26/29, associated with very low serum apoB concentration (p=5.6×10^−4^, **Supplementary Figure S2**), as well as with LDLC (p=9.5×10^−4^), non-HDLC (p=4.8×10^−4^) and TG content in small VLDL particles (p=0.001; **Figure 4, Supplementary Table S7**); the PTVs have not been previously associated with lipid traits. In addition to the *LIPC* PAV association with serum apoA-1 levels described above (p=1.48×10^−7^), the PAVs in *LIPC* were associated with total HDLC and five other lipid phenotypes (p<0.05/19 genes=0.0026; **Figure 4, Supplementary Table S7**). In the *CETP* (MIM:118470) gene, known for genetic disorders of the HDL metabolism, PAVs were associated with serum apoA-1 levels (p=6.9×10^−5^), total HDLC (p=4.0×10^−5^), and seven other lipid measurements in HDL particles, driven by two low-frequency missense variants, rs5880 and rs1800777 previously associated with low HDLC.^42^ PAVs in the hypercholesterolemia-associated *APOE* (MIM:107741) gene were associated with apoB (p=3.5×10^−4^), total HDLC (*p*=8.0×10^−4^), and total cholesterol and cholesterol esters in LDL particles, with large negative effects observed for the previously reported rare p.Glu57Lys (rs201672011) variant.^43^ Finally, the three previously reported PAVs in the *PCSK9* gene, including the protective rs11591147 loss-of-function mutation^44^ were associated with total cholesterol (p=3.0×10^−4^), LDLC (p=0.0014), non-HDLC (p=4.8×10^−4^) and, surprisingly, with serum isoleucine (p=2.9×10^−4^).

**Figure 4:**
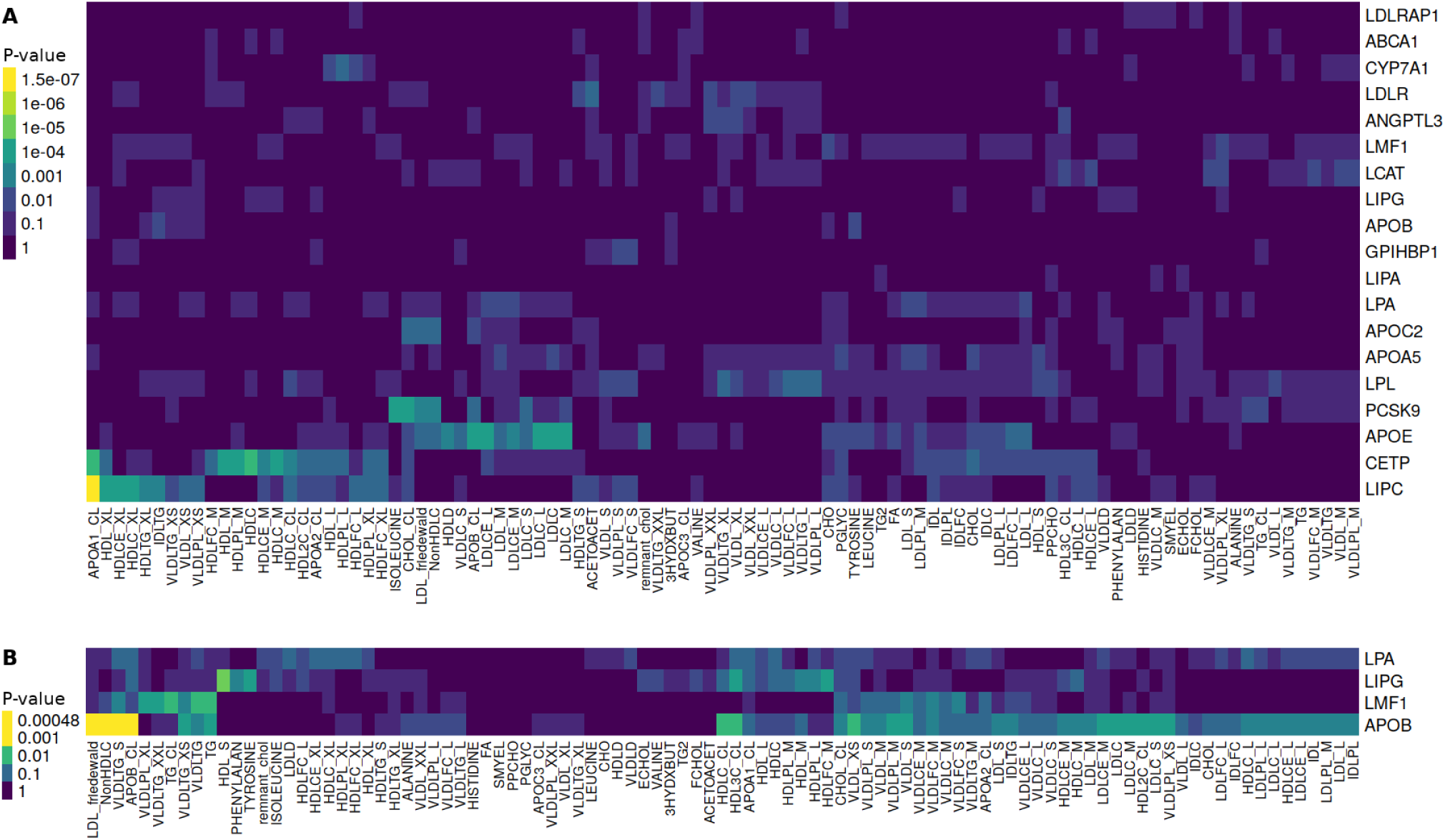
WES-WGS SKAT meta-analysis results across all the studied phenotypes for PAVs (A) and PTVs (B) in genes previously associated with rare lipid disorders. The color indicates the strength of the gene – phenotype association (SKAT p-value).

## Discussion

Dyslipidemia is a considerable risk factor for CVD and DKD.^45^ In addition to the standard clinical lipid laboratory measurements, here we have used apolipoproteins as well as NMR lipid, lipoprotein, and metabolite measurements, combined with WES and WGS to identify genetic variants associated with a total of 97 studied phenotypes. We identified novel associations in genes already implicated in lipid metabolism (e.g. rs113298164 in *LIPC*, two PTVs in *APOB*), as well as multiple novel genes for lipid phenotypes, e.g. *RBM47* and *SBDS* for apoC-III levels, *GTF3C5* for LDLC, and *TRMT5* for phospholipids in VLDL particles.

The lead variant in the single-variant analysis, rs113298164 (*LIPC* p.Thr405Met), was associated with elevated apoA-1 levels (p=7.8×10^−8^). *LIPC* encodes the hepatic lipase, which is the enzyme responsible for TG hydrolysis in IDL particles and, thus, the conversion of IDL to LDL particles. p.Thr405Met is predicted deleterious or probably damaging by SIFT and PolyPhen, and functional studies show that p.Thr405Met reduces hepatic lipase activity.^46, 47^ With 1.7% MAF, it is over 4-fold enriched in the Finnish population. Previously, p.Thr405Met has been identified to cause hepatic lipase deficiency in a compound heterozygous state with another rare mutation in *LIPC*, causing elevated total cholesterol, TG and TG-enriched VLDL and LDL particles, followed by premature atherosclerosis^48^; in our GWAS data, the same rs121912502 (p.Ser301Phe) variant was nominallyassociated (p=0.04) with apoA-1 despite low imputation quality (0.37) and low MAF (0.0002). In addition, three other rare *LIPC* variants (two of them singletons) were found in the WES/WGS data in individuals with markedly higher apoA-1 levels (p-value non-significant) and two of them were predicted deleterious (**Supplementary Table S6**); rs3829462 (p.Phe368Leu) was associated with apoA-1 also in the GWAS data (p=0.02). ApoA-1 is a key structural component of HDL particles – generally associated with lower risk of CVD. While association with higher apoA-1 and HDLC may seem contradictory to the association with high total cholesterol and hypertriglyceridemia, severe hepatic lipase deficiency is characterized by an increase in apoA-1, HDLC, and HDL TG content^49^, all seen in our data as well.

Common variants in the *LIPC* gene are strongly associated with serum HDLC and apoA-1 levels.^12^ In a recent Mendelian Randomization analysis, variants associated with elevated apoA-1 levels were associated with lower risk of CAD in the univariate analysis; however, this effect disappeared when accounted for variants affecting apoB levels.^12^ Altogether, these findings suggest that rare *LIPC* variants, rs113298164 (p.Thr405Met) in particular, affect hepatic lipase activity leading to elevated apoA-1 levels, but likely without direct consequence to CVD risk.

Importantly, we identified two PTVs in *APOB* associated with drastically low serum apoB concentrations (**Supplementary Figure 2**); to our knowledge, these variants have not been previously associated with lipid traits. However, with only three individuals, we do not see any association with CVD endpoints.

In SKAT-O gene burden tests, we showed that *RBM47* was associated with lower apoC-III levels. This association was driven by rs564837143, a 21 bp inframe deletion found in the WGS data, located in the 6th exon. Even though the association at rs564837143 (Ala496-Ala502del) did not replicate in our GWAS data, we obtained external validation, as the burden of rare deleterious variants in *RBM47* was associated with TG-to-HDLC ratio (p=0.0093) in WES of 20,917 individuals.^38^ Furthermore, another rare missense variant in the 7th exon was recently shown to have a large impact on blood pressure in a large meta-analysis.^50^ *RBM47* encodes an RNA-binding protein essential for post-transcriptional modification of the apoB mRNA in particular. This modification creates a premature stop codon in the transcript, resulting in the production of the shorter intestinal isoform apoB-48 instead of the longer isoform apoB-100 produced by the liver.^51^ Of note, we have previously shown that apoB-48 is elevated in individuals with type 1 diabetes both at fasting and postprandially.^52^ In this study, however, the identified *RBM47* deletion was associated with lower serum apoC-III levels. Whereas one copy of apoB is firmly embedded within the surface of each TRL (i.e., chylomicrons, VLDL, and IDL) and LDL particle, apoC-III is dynamically redistributed between these and HDL particles in the circulation.^53^ ApoC-III is an important regulator of TG metabolism that, through multiple pathways, impairs the clearance of the atherosclerotic, apoB-containing TRLs and their remnants. One key action of apoC-III is the inhibition of lipoprotein lipase, and to some extent, also hepatic lipase encoded by the *LIPC* gene.^54^ There is increasing evidence – also from genetic studies of a rare *APOC3* loss-of-function variant^55, 56^ – that apoC-III is an independent cardiovascular risk factor, and clinical trials on apoC-III lowering therapies have yielded positive results in those with high TG. ApoC-III is an important CVD risk factor also in individuals with type 1 diabetes^57^. In contrast to that, the *RBM47* Ala496-Ala502 carriers with lower serum apoC-III concentrations were not significantly protected from CVD in our data (**Figure 2E**); however, the low number of observed carriers does not allow us to draw definite conclusions.

PAVs in *GTF3C5* were associated with total cholesterol, LDLC, and non-HDLC. Among the eight PAVs, six were predicted deleterious by SIFT and/or PolyPhen. One of them, chr9:133042147_C/T (p.His72Tyr), is a novel variant, with one heterozygous carrier found in our data (verified as good quality from the aligned BAM-file). Another variant, rs189383196, is either a high impact start-lost variant, or a missense variant (p.Met126Thr), depending on the transcript, with over 80-fold enrichment in Finns. The association for the strongest individual variant, rs202207045 (p.Ala382Val), was also replicated in the GWAS data (p=0.02 for LDLC and non-HDLC). The PAVs in this gene were associated with multiple circulatory phenotypes, e.g. non-ischemic cardiomyopathy (p=2.8×10^−5^) in the independent FinnGen data. Interestingly, the strongest association within the *GTF3C5* region in the FinnGen GWAS data was at rs671412, 28 kbp downstream, with statin medication (p=3.4×10^−7^). *GTF3C5* encodes a DNA-binding general transcription factor IIIC subunit 5, expressed in all tissues, and little is known about the function of this gene. PAVs in *TRMT5* were associated with phospholipids in extra small VLDL particles, both in WES and WGS separately, as well as in WES-WGS SKAT-O meta-analysis. Among the eight variants, five were predicted deleterious. As supporting evidence, the deleterious missense variant with the strongest association with lower phospholipids in VLDL particles, was associated with higher risk of stroke in the FinnGen data (p=1.90×10^−4^). *TRMT5* encodes a tRNA methyltransferase 5 involved in mitochondrial tRNA methylation.

Other novel findings worth mentioning are PTVs in *SBDS*, as well as PAVs in *CYP3A43, PTGER3*, and *AKAP3*. Loss-of-function variants in S*BDS* cause autosomal recessive Shwachman-Diamond Syndrome 1, characterized by exocrine pancreatic dysfunction among other symptoms^58^; our observed association between heterozygous *SBDS* PTVs and apoC-III may be affected by a similar pathway. PAVs in *CYP3A43* were associated with LDL cholesterol esters in WGS and replicated in WES; *CYP3A43* was the only gene with evidence of association with clinical outcome in our WES-WGS data (SKAT p=0.004 for DKD, rank 43/17,578 genes). While little is known about the gene, it encodes one of the cytochrome P450 proteins, which are involved in the synthesis of cholesterol, steroids and other lipids and, importantly, metabolize most of the drugs and can cause toxic drug-drug interactions, e.g. with the statins.^59^

This study represents the first comprehensive analysis of PAVs associated with detailed lipid, apolipoprotein, and lipoprotein phenotypes in individuals with type 1 diabetes. While some of the observed associations may be specific to individuals with diabetes e.g. through disturbances in insulin signaling, we hypothesize that many of the associations observed in this high risk population may be generalized to the wider population, as was seen in our replication attempts both for the single variant and gene level findings in the general population data sets. Despite a limited number of samples, we were able to identify multiple novel genetic associations. Of note, many of the identified variants were markedly enriched in the Finnish population, e.g. the 80-fold Finnish-enriched *GTF3C5* PAVs, providing one potential explanation why these variants have not been detected in earlier studies. It is of note that many previous, larger studies were either based on chip genotyping,^60^ or included only the standard clinical lipid measurements such as total cholesterol, LDLC, HDLC and TG levels (quantitative trait analysis in a subset of *Flannick et al*.).^40^ While limited evidence of replication was found for the single-variant associations in the FinnDiane GWAS data, many of the identified PAVs or genes were associated with relevant metabolic traits and clinical endpoints in larger external data sets.

Previous studies suggest that apoC-III is an important, independent risk factor for CVD. While we identified a seven amino acid deletion in *RBM47* associated with lower apoC-III levels, the variant was not associated with CVD in longitudinal analysis, and further studies are needed to elucidate the biological mechanism that it exerts on the apolipoprotein levels. To conclude, in addition to the previously known associations, we identified multiple novel genes involved in lipid metabolism.

## Data Availability

The sequencing data supporting the current study have not been deposited in a public repository because of restrictions due to the study consent and local regulations.

## Supplemental Information description

Supplemental material (.docx), including Tables S1-S7 and Figures S1 and S2.

## Acknowledgements

We want to acknowledge the participants and investigators of the FinnGen study for the look-ups of the lead findings; and ELIXIR Finland node hosted at CSC – IT Center for Science for ICT resources that enabled the WES and WGS data processing. We further acknowledge the physicians and nurses at each FinnDiane center participating in the collection of the patient data.

## Funding

The study was supported by grants from Folkhälsan Research Foundation, Wilhelm and Else Stockmann Foundation, “Liv och Hälsa” Society, Sigrid Juselius Foundation, Helsinki University Central Hospital Research Funds (EVO TYH2018207), Novo Nordisk Foundation (NNF OC0013659), Academy of Finland (299200, and 316664), European Foundation for the Study of Diabetes (EFSD) Young Investigator Research Award funds, and an EFSD award supported by EFSD/Sanofi European Diabetes Research Programme in Macrovascular Complications.

## Declaration of Interests

P-H G has received investigator-initiated research grants from Eli Lilly and Roche, is an advisory board member for AbbVie, Astellas, AstraZeneca, Bayer, Boehringer Ingelheim, Cebix, Eli Lilly, Janssen, Medscape, Merck Sharp & Dohme, Mundipharma, Nestlé, Novartis, Novo Nordisk and Sanofi; and has received lecture fees from AstraZeneca, Boehringer Ingelheim, Eli Lilly, Elo Water, Genzyme, Merck Sharp & Dohme, Medscape, Novartis, Novo Nordisk, PeerVoice and Sanofi. Other authors declare no competing interests.

## Web Resources

AMP T2D-GENES exome sequencing, https://t2d.hugeamp.org/

Beagle, https://faculty.washington.edu/browning/beagle/b4_1.html

dbSNP, https://www.ncbi.nlm.nih.gov/snp/

Eagle, https://data.broadinstitute.org/alkesgroup/Eagle/

FinnGen study GWAS results, freeze 3: https://r3.finngen.fi/

FinnGen study GWAS results, freeze 4: https://r4.finngen.fi/

GATK, https://gatk.broadinstitute.org/hc/en-us

Human Protein Atlas, https://www.proteinatlas.org/

OMIM, https://www.omim.org/

Primer3, https://primer3.org/

R survival package: https://cran.r-project.org/web/packages/survival/

RAREMETAL, https://genome.sph.umich.edu/wiki/RAREMETAL_Documentation

Rvtests, http://zhanxw.github.io/rvtests/

SnpEff, https://pcingola.github.io/SnpEff/

UCSC LiftOver program, https://genome-store.ucsc.edu/

Variant effect predictor VEP, https://www.ensembl.org/Tools/VEP

## Notes

### Author Declarations

The study protocol was approved by the Ethical Committee of the Helsinki and Uusimaa Hospital District and local ethics committees and participants gave their informed consent before recruitment. This study was performed following the Declaration of Helsinki.

