## Supplementary Material for "Novel protein-altering variants associated with serum apolipoprotein and lipid levels"

**Supplemental material**

### Supplementary Table S1. Lipid, lipoprotein, apolipoprotein, and metabolite phenotypes.

| Phenotype | Abbreviation | N WES | N WGS | Method |
|---|---|---|---|---|
| Serum apolipoprotein A-I [mg/dl] | APOA1 | 469 | 449 | Cobas Mira analyzer (Boehringer-Mannheim) until January 2002, Wako Chemicals GmbH (Neuss, Germany) until January 2006, Konelab 60i analyser and commercial kit (Thermo Fischer Scientific, Waltham, MA, USA) from 2006 onward. |
| Serum apolipoprotein A-II [mg/dl] | APOA2 | 455 | 383 | Cobas Mira analyzer using immunoprecipitation (Boehringer-Mannheim) until August 2001, and thereafter with a polyclonal antibody. |
| Serum apolipoprotein B-100 [mg/dl] | APOB | 469 | 449 | Immunoassay (Orion Diagnostica, Espoo, Finland) until January 2006, and since then with Konelab 60i analyser and a commercial kit (Thermo Fischer Scientific, Waltham, MA, USA). |
| Serum Apolipoprotein C-III [mg/dl] | APOC3 | 323 | 294 | Immunoassay (Kamiya Biomedical Company, Tukwila, WA 98168, USA). |
| Serum total cholesterol [mmol/l] | CHOL CL | 469 | 451 | Autoanalyzer with an enzymatic technique (Hoffman-La Roche, Basel, Switzerland until November 2001; ABX Diagnostics, Montpellier, France until January 2006; and Konelab 60i analyser and a commercial kit (Thermo Fischer Scientific, Waltham, MA, USA) from this point onward. |
| Serum HDL cholesterol [mmol/l] | HDLC | 469 | 450 | Enzymatic method, described in Tolonen, Forsblom et al. 2008. |
| Serum $HDL_2$ cholesterol [mmol/l] | HDL2C | 463 | 441 | Calculated by subtracting of HDL3-cholesterol from the total HDL-cholesterol. |
| Serum $HDL_3$ cholesterol [mmol/l] | HDL3C | 463 | 441 | Enzymatic method, described in Tolonen, Forsblom et al 2008. |
| Serum triglycerides [mmol/l] | TG | 469 | 451 | Autoanalyzer with an enzymatic technique (Hoffman-La Roche, Basel, Switzerland until November 2001; ABX Diagnostics, Montpellier, France until January 2006; and Konelab 60i analyser and a commercial kit (Thermo Fischer Scientific, Waltham, MA, USA) from this point onward. |
| LDL cholesterol [mmol/l] calculated with Friedewald equation, TotalChol - (Triglyceride / 5) - HDL | LDL friedewald | 469 | 451 | Friedewald equation described in Friedewald, Levy et al. 1972. |
| Non-HDL cholesterol (TOTCHOL-HDLCHOL) [mmol/l] | NonHDLC | 469 | 450 | Calculated by subtracting of serum HDL-cholesterol from the serum total cholesterol. |
| Remnant cholesterol, TotalChol - HDL - LDL | Remnant chol | 469 | 450 | Calculated by subtracting of HDL-and LDL-cholesterol from the serum total cholesterol described in Varbo, benn et al. 2014. |
| Total cholesterol | CHOL | 419 | 329 | NMR |
| Total triglycerides | TG | 419 | 329 | NMR |
| VLDL particle diameter | VLDLD | 115 | 84 | NMR |
| LDL particle diameter | LDLD | 115 | 84 | NMR |
| HDL particle diameter | HDLD | 115 | 84 | NMR |
| Total lipids in extremely large VLDL | VLDL XXL | 419 | 329 | NMR |
| Phospholipid in extremely large VLDL | VLDLPL XXL | 419 | 329 | NMR |
| Triglycerides in extremely large VLDL | VLDLTG XXL | 304 | 245 | NMR |
| Total lipids in extra large VLDL | VLDL XL | 419 | 329 | NMR |
| Phospholipid in extra large VLDL | VLDLPL XL | 258 | 187 | NMR |
| Triglycerides in extra large VLDL | VLDLTG XL | 419 | 319 | NMR |
| Total lipids in large VLDL | VLDL L | 419 | 319 | NMR |

| | | | | |
|---|---|---|---|---|
| Cholesterol in large VLDL | VLDLC L | 419 | 319 | NMR |
| Cholesterol ester in large VLDL | VLDLCE L | 419 | 319 | NMR |
| Free cholesterol in large VLDL | VLDLFC L | 419 | 319 | NMR |
| Phospholipid in large VLDL | VLDLPL L | 419 | 319 | NMR |
| Triglycerides in large VLDL | VLDLTG L | 419 | 319 | NMR |
| Total lipids in medium VLDL | VLDL M | 419 | 319 | NMR |
| Cholesterol in medium VLDL | VLDLC M | 419 | 319 | NMR |
| Cholesterol ester in medium VLDL | VLDLCE M | 419 | 319 | NMR |
| Free cholesterol in medium VLDL | VLDLFC M | 419 | 319 | NMR |
| Phospholipid in medium VLDL | VLDLPL M | 419 | 319 | NMR |
| Triglycerides in medium VLDL | VLDLTG M | 419 | 319 | NMR |
| Total lipids in small VLDL | VLDL | 419 | 319 | NMR |
| Cholesterol in small VLDL | VLDLC | 419 | 319 | NMR |
| Free cholesterol in small VLDL | VLDLFC | 419 | 319 | NMR |
| Phospholipid in small VLDL | VLDLPL | 419 | 319 | NMR |
| Triglycerides in small VLDL | VLDLTG | 419 | 319 | NMR |
| Total lipids in extra small VLDL | VLDL XS | 419 | 319 | NMR |
| Phospholipid in extra small VLDL | VLDLPL XS | 419 | 319 | NMR |
| Triglycerides in extra small VLDL | VLDLTG XS | 419 | 319 | NMR |
| Triglycerides in VLDL particles | VLDLTG | 419 | 319 | NMR |
| Total lipid in IDL particles | IDL | 419 | 319 | NMR |
| Cholesterol in IDL particles | IDLC | 419 | 319 | NMR |
| Free cholesterol in IDL particles | IDLFC | 419 | 319 | NMR |
| Phospholipid in IDL particles | IDLPL | 419 | 319 | NMR |
| Triglyceride in IDL particles | IDLTG | 419 | 319 | NMR |
| Total lipid in large LDL | LDL L | 419 | 319 | NMR |
| Cholesterol in large LDL | LDLC L | 419 | 319 | NMR |
| Cholesterol ester in large LDL | LDLCE L | 419 | 319 | NMR |
| Free cholesterol in large LDL | LDLFC L | 419 | 319 | NMR |
| Phospholipid in large LDL | LDLPL L | 419 | 319 | NMR |
| Total lipid in medium LDL | LDL M | 419 | 319 | NMR |
| Cholesterol in medium LDL | LDLC M | 419 | 319 | NMR |
| Cholesterol ester in medium LDL | LDLCE M | 419 | 319 | NMR |
| Phospholipid in medium LDL | LDLPL M | 419 | 319 | NMR |
| Total lipid in small LDL | LDL S | 419 | 319 | NMR |
| Cholesterol in small LDL | LDLC S | 419 | 319 | NMR |
| Cholesterol in LDL particles | LDLC | 419 | 319 | NMR |
| Total lipid in extra large HDL | HDL XL | 419 | 319 | NMR |
| Cholesterol in extra large HDL | HDLC XL | 419 | 319 | NMR |

| | | | | |
|---|---|---|---|---|
| Cholesterol ester in extra large HDL | HDLCE XL | 419 | 319 | NMR |
| Free cholesterol in extra large HDL | HDLFC XL | 419 | 319 | NMR |
| Phospholipid in extra large HDL | HDLPL XL | 419 | 319 | NMR |
| Triglyceride in extra large HDL | HDLTG XL | 419 | 319 | NMR |
| Total lipid in large HDL | HDL L | 419 | 319 | NMR |
| Cholesterol in large HDL | HDLC L | 419 | 319 | NMR |
| Cholesterol ester in large HDL | HDLCE L | 419 | 319 | NMR |
| Free cholesterol in large HDL | HDLFC L | 419 | 319 | NMR |
| Phospholipid in large HDL | HDLPL L | 419 | 319 | NMR |
| Total lipid in medium HDL | HDL M | 419 | 319 | NMR |
| Cholesterol in medium HDL | HDLC M | 419 | 319 | NMR |
| Cholesterol ester in medium HDL | HDLCE M | 419 | 319 | NMR |
| Free cholesterol in medium HDL | HDLFC M | 419 | 319 | NMR |
| Phospholipid in medium HDL | HDLPL M | 419 | 319 | NMR |
| Total lipid in small HDL | HDL S | 419 | 319 | NMR |
| Triglycerides in small HDL | HDLTG S | 419 | 319 | NMR |
| Cholesterol in HDL particles | HDLC | 419 | 319 | NMR |
| Total fatty acids (mmol/l) | FA | 158 | 118 | NMR |
| Free cholesterol (mmol/l) | FCHOL | 158 | 118 | NMR |
| Total triglycerides (mmol/l) | TG2 | 155 | 115 | NMR |
| Esterified cholesterol (mmol/l) | ECHOL | 158 | 118 | NMR |
| Total phophoglycerides (mmol/l) | PGLYC | 143 | 110 | NMR |
| Phosphatidylcholine and other cholines (mmol/l) | PPCHO | 143 | 110 | NMR |
| Sphingomyelins (mmol/l) | SMYEL | 132 | 106 | NMR |
| Total cholines and other N-trimethyl compounds (mmol/l) | CHO | 122 | 91 | NMR |
| Histidine | HISTIDINE | 112 | 87 | NMR |
| Isoleucine | ISOLEUCINE | 419 | 346 | NMR |
| Leucine | LEUCINE | 116 | 88 | NMR |
| Phenylalanine | PHENYLALAN | 115 | 88 | NMR |
| Tyrosine | TYROSINE | 114 | 88 | NMR |
| Valine | VALINE | 419 | 345 | NMR |
| Alanine | ALANINE | 419 | 346 | NMR |
| 3-hydroxybutyrate | 3HYDXBUT | 418 | 346 | NMR |
| Acetoacetate | ACETOACET | 417 | 346 | NMR |

N: Number of patients in WES and WGS cohorts. NMR: Nuclear Magnetic Resonance.

**Supplementary Table S2: Number of PAVs and PTVs in the WES and WGS data. Counts are calculated for the total cholesterol lab measurements.**

| | | PAV | | | PTV | | |
|---|---|---|---|---|---|---|---|
| | N | MAF=50% | MAF=5% | MAF=1% | MAF=50% | MAF=5% | MAF=1% |
| **WGS** | 451 | 101,718 | 80,700 | 65,534 | 9,577 | 7,866 | 6,497 |
| **WES** | 469 | 42,682 | 34,881 | 27,257 | 2,240 | 2,009 | 1,749 |
| **meta** | 920 | 115,923 | 97,159 | 82,428 | 7,554 | | |
| | | | **proportion of PAV** | | **proportion of PAV** | **Proportion of PTV** | |
| **WGS** | | | 79 % | 64 % | 9 % | 82 % | 68 % |
| **WES** | | | 82 % | 64 % | 5 % | 90 % | 78 % |
| **meta** | | | 84 % | 71 % | 7 % | | |

PTV= Protein-truncating variant. PAV= Protein-altering variant.

**Supplementary Table S3: Single variant WES+WGS meta-analysis results for associations with $p<1\times10^{-5}$**

| Gene | Variant | MAF | Phenotype | P | Effect | Dir | P GWAS |
|---|---|---|---|---|---|---|---|
| *CAPZB* | 1:19448881:G:A | 0.013 | TG | $7.2 \times10^{-6}$ | Benign, | ?- | 0.41 |
| | (rs79308175,missense) | | VLDLPL M | $5.9\times10^{-6}$ | Possibly deleterious | ?- | 0.33 |
| | | | VLDLTG XS | $7.2\times10^{-6}$ | | ?- | 0.32 |
| | | | VLDLTG S | $4.0\times10^{-6}$ | | ?- | 0.41 |
| *PGLYRP4* | 1:153341637:G:T | 0.002 | HDLC | $8.7 \times10^{-6}$ | Benign, | -- | 0.76 |
| | (rs73014502,missense) | | | | Tolerated | | |
| *SERPINI2* | 3:167467090:T:C | 0.467 | HDLD | $2.9 \times10^{-6}$ | Possibly damaging | ++ | 0.051 |
| | (rs9841174, missense) | | | | Deleterious | | |
| *RBM47* | 4:40432687: | 0.005 | APOC3 | $2.5 \times10^{-6}$ | unknown | ?- | 0.08 |
| | AGCGGCTGCGGCGGCTGCGGCC:A | | | | | | |
| *NOCT* | 4:139045355:G:A | 0.005 | Total Cholesterol | $3.8 \times10^{-6}$ | Benign, | ++ | 0.63 |
| | (rs144899070, missense) | | | | Tolerated | | |
| *PPIC* | 5:123023945:T:C | 0.156 | HDLFC L | $5.0 \times10^{-6}$ | Benign | ?+ | 0.87 |
| | (rs451195, missense) | | | | Deleterious | | |
| *CFAP206* | 6: 87415755:A:G | 0.005 | HDLC | $3.6 \times10^{-6}$ | Probably damaging, | ?+ | 0.99 |
| | (rs35978098, missense) | | | | Tolerated | | |
| *SBDS* | 7:66994210:A:G | 0.005 | APOC3 | $5.9 \times10^{-6}$ | Pathogenic (ClinVar) | ?+ | 0.19 |
| | (rs113993993, splice donor) | | | | | | |
| *DEFT1P* | 8:6989658: C:T | 0.020 | VLDLPL XL | $1.2 \times10^{-6}$ | Unknown | ?+ | - |
| | (rs797006828, splice-donor) | | | | | | |
| *NRG1* | 8:32754452:G:T | 0.015 | LDLPL M | $9.0 \times10^{-6}$ | Possibly damaging, | ++ | 0.62 |
| | (rs74942016, missense) | | | | Deleterious | | |
| *IFNW1* | 9:21141554:G:C | 0.005 | HDLTG XL | $3.7 \times10^{-6}$ | Benign, | ++ | 0.43 |
| | (rs201154486, missense) | | | | Deleterious | | |
| *GTF3C5* | 9:133054787:C:T | 0.007 | LDL Friedewald, | $1.3 \times10^{-6}$ | Benign, | -- | **0.02 (-)** |
| | (rs202207045, missense) | | Non-HDLC | $7.3 \times10^{-7}$ | Tolerated | -- | **0.02 (-)** |
| | | | Total Cholesterol | $1.2 \times10^{-6}$ | | -- | 0.11 |
| *TET1* | 10:68572915:T:A | 0.091 | LDL M | $8.3 \times10^{-6}$ | Benign, | ?- | 0.57 |
| | (rs12773594, missense) | | | | Tolerated | | |
| *ANO9* | 11:433867:G:A | 0.185 | HDLC | $5.7 \times10^{-6}$ | Benign, Tolerated | ++ | 0.33 |
| | (rs12575508, missense) | | | | | | |
| *OVCH1* | 12:29464617:G:A | 0.191 | Remnant | $2.9 \times10^{-6}$ | Possibly damaging, | ++ | 0.83 |
| | (rs11050243, missense) | | | | Deleterious | | |
| *FNDC3A* | 13:49201861:A:G | 0.064 | CHOL CL | 8.8 ×10-$^{6}$ | Benign, | ++ | **0.04 (+)** |
| | (rs45604939, missense) | | HDLPL M | $5.2 \times10^{-6}$ | Deleterious | ++ | 0.03 (-) |
| | | | LDLPL L | $5.7 \times10^{-6}$ | | ++ | 0.64 |
| | | | VLDL S | $8.0 \times10^{-6}$ | | ++ | 0.22 |
| | | | VLDL XS | $6.1 \times10^{-6}$ | | ++ | 0.45 |
| | | | VLDLFC S | $7.0 \times10^{-6}$ | | ++ | 0.40 |
| *SYNE3* | 14:95417986:G:A | 0.650 | IDLC | $9.0 \times10^{-6}$ | Benign, | ++ | 0.64 |
| | (rs12434757,missense) | | | | Tolerated | | |
| *GABRG3* | 15:27271593: GAGTC:G | 0.299 | VLDL XL | $4.9 \times10^{-6}$ | Unknown | ?- | 0.19 |
| | (5' UTR variant in 2 alternative transcripts) | | VLDLTG XL | $4.1 \times10^{-6}$ | | ?- | 0.21 |
| *LIPC* | 15:58563549:C:T | 0.017 | APOA1 | $7.8 \times10^{-8}$ | Probably damaging | ++ | 0.89 |
| | (rs113298164, missense) | | | | Deleterious | | |
| *METTL16* | 17: 2475244:T:G | 0.068 | LDLPL L | $8.2 \times10^{-6}$ | Benign, | ?- | 0.96 |
| | (rs2028600, missense) | | | | Tolerated | | |
| *MARCH10* | 17:62736189:G:A | 0.003 | IDLFC | $9.3 \times10^{-6}$ | Possibly damaging, | -- | 0.90 |
| | (rs147046907, missense) | | IDLPL | $7.4 \times10^{-6}$ | Deleterious | -- | 1.00 |
| | | | LDLPL M | $7.5 \times10^{-6}$ | | -- | 0.63 |
| | | | VLDL M | $6.9 \times10^{-6}$ | | -- | 0.73 |
| | | | VLDLFC M | $1.2 \times10^{-6}$ | | -- | 0.67 |
| | | | VLDLPL M | $1.6 \times10^{-6}$ | | -- | 0.59 |
| | | | VLDLPL_XS | $1.1 \times10^{-6}$ | | -- | 0.76 |
| *MYO15B* | 17:75591977:G:T | 0.252 | HDLTG S | $1.4\times10^{-6}$ | Possibly damaging | ++ | 0.95 |
| | (rs736522, missense) | | | | Tolerated | | |
| *OR7A10* | 19:14841426:A:G | 0.149 | Tyrosine | $3.7 \times10^{-6}$ | Benign, | ++ | 0.77 |
| | (rs12972670, missense) | | | | Tolerated | | |
| *ZNF274* | 19: 58206912:A:G | 0.021 | VLDL L | $9.3 \times10^{-6}$ | Benign, | ?- | 0.87 |
| | (rs45580533, missense) | | VLDLTG L | $5.5 \times10^{-6}$ | Tolerated | | 0.81 |
| *LBP* | 20:38346542:C:T | 0.052 | LDL Friedewald | $4.9 \times10^{-6}$ | Benign, | ++ | 0.49 |
| | (rs2232580, missense) | | | | Tolerated | | |

| Gene | Variant | MAF | Phenotype | P | Effect | Dir | P GWAS |
|---|---|---|---|---|---|---|---|
| *AP000233.3* | 21:25135053:C:T (rs2829553, splice-donor) | 0.223 | APOA1 | $2.1 \times 10^{-6}$ | Low confidence pLoF | ?- | 0.25 |
| *KRTAP13-1* | 21:30396529:G:A (rs151147550,missense) | 0.007 | APOC3 | $4.9 \times 10^{-6}$ | Benign, Tolerated | ++ | 0.76 |

MAF: Minor allele frequency, Effect: SIFT and Polyphen-2 predictions of the effect of the variant. P SKAT/SKATO: P-value of the SKAT or SKAT-O association test. Dir: Effect direction (for ALT allele) in WES and WGS, respectively. P GWAS: P-value from GWAS analysis, with direction of effect (+/-) indicated for associations with p-value <0.05. Associations with p-value < 0.05 and consistent direction of effect are highlighted with bold.

**Supplementary Table S4: WES-WGS SKAT meta-analysis on CVD, CAD, Strokes, and DKD for the lead genes.**

| GENE | N VAR | AVG_AF | P CVD | P CAD | P Stroke | P DKD |
|---|---|---|---|---|---|---|
| **PAV** | | | | | | |
| *RBM47* | 4 | 0.002 | 0.444 | 1.000 | 0.148 | 0.332 |
| *CYP3A43* | 7 | 0.005 | 0.360 | 0.091 | 0.675 | **0.004** |
| *GTF3C5* | 9 | 0.002 | 0.066 | 0.270 | 0.465 | 0.844 |
| *AKAP3* | 10 | 0.015 | 0.733 | 0.599 | 0.827 | 0.822 |
| *TRMT5* | 8 | 0.003 | 0.307 | 0.567 | 0.610 | 0.736 |
| *LIPC* | 6 | 0.017 | 0.325 | 0.726 | 0.508 | 0.658 |
| **PTV** | | | | | | |
| *PTGER3* | 1 | 0.010 | 0.672 | 0.278 | 0.157 | 0.476 |
| *SBDS* | 3 | 0.002 | 0.595 | 0.276 | 0.536 | 0.061 |
| *DEFT1P/ DEFT1P2* | 1 | 0.024 | 0.448 | 0.197 | 0.707 | 0.265 |

N Var: number of PAV/ PTV variants with MAF ≤ 5% within the gene. AVG_AF: average allele frequency of the variants. P CVD/ CAD/ Stroke/ DKD: WES-WGS SKAT meta-analysis P-value.

**Supplementary Table S5: Single variant score test association results for PTVs and PAVs within the lead genes from the SKAT WES+WGS meta-analysis.**

| | WES+WGS SKAT meta-analysis | | | | WES+WGS meta | | | GWAS | | | |
|---|---|---|---|---|---|---|---|---|---|---|---|
| **GENE** | **Type** | **Pheno** | **P_LIU** | **Variants** | **MAF** | **EFFECT** | **P-values** | **N** | **AF** | **Effect** | **P** |
| *DEFT1P/ DEFT1P2* | PTV | VLDLPL XL | 1.23E-06 | 8:6989658:C:T (rs797006828, splice donor variant ) | 0.020 | 1.157 | **1.23E-06** | | | | |
| *SBDS* | PTV | apoC-III | 5.37E-06 | 7:66994210:A:G (rs113993993, splice donor variant); | 0.005 | 1.865 | **5.86E-06** | 2982 | 0.008 | 0.22 | 0.19 |
| | | | | 7:66994286:T:A (rs113993991, stop gain) | 0.001 | 0.603 | 0.55 | | | | |
| *LIPC* | PAV | apoA1 | 1.48E-07 | 15:58541794:G:A (rs6078*); | 0.032 | 0.171 | 0.20 | 3759 | 0.058 | 0.04 | 0.41 |
| | | | | 15:58541843:C:T (rs562988299*); | 0.001 | 0.877 | 0.38 | | | | |
| | | | | 15:58541944:G:A (rs201563586*); | 0.001 | 1.794 | 0.07 | 3759 | 0.002 | 0.12 | 0.67 |
| | | | | 15:58548387:C:T (rs121912502*); | 0.001 | 0.937 | 0.35 | 3759 | 0.0002 | 2.56 | **0.04**** |
| | | | | 15:58560880:A:C (rs3829462*); | 0.045 | 0.001 | 0.99 | 3759 | 0.046 | 0.12 | **0.02** |
| | | | | 15:58563549:C:T (rs113298164*) | 0.017 | 0.980 | **7.77E-08** | 3759 | 0.058 | 0.01 | 0.89 |
| *GTF3C5* | PAV | Non-HDLC | 6.86E-07 | 9:133042147:C:T (-) ; | 0.001 | 0.596 | 0.55 | | | | |
| | | | | 9:133043731:T:C (rs189383196, start lost, missense); | 0.004 | 0.311 | 0.38 | 4636 | 0.010 | 0.14 | 0.17 |
| | | | | 9:133043838:C:A (rs150056568)*; | 0.001 | 0.017 | 0.98 | 4636 | 0.003 | 0.18 | 0.33 |
| | | | | 9:133054459:A:G (rs369889499*); | 0.001 | -0.392 | 0.58 | 4636 | 0.001 | -0.08 | 0.84 |
| | | | | 9:133054733:A:G (rs1302997790)*; | 0.001 | 0.840 | 0.40 | | | | |
| | | | | 9:133054787:C:T (rs202207045*); | 0.007 | -1.382 | **7.26E-07** | 4636 | 0.008 | -0.28 | **0.02** |
| | | | | 9:133056848:G:A (rs637435)*; | 0.001 | -0.596 | 0.55 | | | | |
| | | | | 9:133057819:T:C (rs111893665)* | 0.001 | -0.120 | 0.90 | 4636 | 0.001 | 0.17 | 0.60 |
| *GTF3C5* | PAV | LDL Friedewald | 9.27E-07 | 9:133042147:C:T (-) ; | 0.001 | 0.642 | 0.52 | | | | |
| | | | | 9:133043731:T:C (rs189383196, start lost, missense); | 0.003 | -0.309 | 0.45 | 4584 | 0.010 | 0.16 | 0.11 |
| | | | | 9:133043838:C:A (rs150056568)*; | 0.001 | 0.100 | 0.89 | 4584 | 0.003 | 0.19 | 0.31 |
| | | | | 9:133054459:A:G (rs369889499*); | 0.001 | -0.302 | 0.67 | 4584 | 0.001 | -0.01 | 0.99 |
| | | | | 9:133054733:A:G (rs1302997790)*; | 0.001 | 0.956 | 0.34 | | | | |
| | | | | 9:133054787:C:T (rs202207045*); | 0.007 | -1.350 | **1.31E-06** | 4584 | 0.008 | -0.28 | **0.02** |
| | | | | 9:133056848:G:A (rs637435)*; | 0.001 | -0.690 | 0.49 | | | | |
| | | | | 9:133057819:T:C (rs111893665)* | 0.001 | -0.131 | 0.90 | 4584 | 0.001 | 0.07 | 0.83 |
| *GTF3C5* | PAV | CHOL CL | 1.30E-06 | 9:133042147:C:T (-) ; | 0.001 | 1.113 | 0.27 | | | | |
| | | | | 9:133043731:T:C (rs189383196, start lost, missense); | 0.004 | 0.253 | 0.48 | 4653 | 0.010 | 0.12 | 0.23 |
| | | | | 9:133043838:C:A (rs150056568)*; | 0.001 | -0.003 | 1.00 | 4653 | 0.003 | 0.18 | 0.35 |
| | | | | 9:133054459:A:G (rs369889499*); | 0.001 | -0.344 | 0.63 | 4653 | 0.001 | -0.10 | 0.78 |
| | | | | 9:133054733:A:G (rs1302997790)*; | 0.001 | 0.671 | 0.50 | | | | |
| | | | | 9:133054787:C:T (rs202207045*); | 0.007 | -1.353 | **1.23E-06** | 4653 | 0.008 | -0.18 | 0.11 |
| | | | | 9:133056848:G:A (rs637435)*; | 0.001 | -0.885 | 0.38 | | | | |
| | | | | 9:133057819:T:C (rs111893665)* | 0.001 | 0.248 | 0.80 | 4653 | 0.001 | 0.31 | 0.32 |
| *TRMT5* | PAV | VLDLPL XS | 7.87E-07 | 14:60975180:G:A (rs147405788*); | 0.001 | -1.575 | 0.03 | 1466 | 0.002 | -0.08 | 0.82 |

| | | | | | | | | | | | |
|---|---|---|---|---|---|---|---|---|---|---|---|
| | | | | 14:60975182:T:C (-)*; | 0.001 | -1.386 | 0.17 | | | | |
| | | | | 14:60975636:G:A (rs45604437*); | 0.007 | -1.094 | **5.76E-04** | 1466 | 0.003 | -0.23 | 0.53 |
| | | | | 14:60979242:T:C (rs138139551*); | 0.002 | -0.599 | 0.30 | 1466 | 0.003 | -0.04 | 0.91 |
| | | | | 14:60979257:T:C (rs191207997*); | 0.001 | -0.713 | 0.48 | 1466 | 0.001 | -0.36 | 0.53 |
| | | | | 14:60979303:C:G (rs1285433520)*; | 0.001 | 0.400 | 0.69 | | | | |
| | | | | 14:60979401:G:A (rs746471581*); | 0.001 | 1.362 | 0.17 | | | | |
| | | | | 14:60979429:T:A (rs115400838)* | 0.007 | -1.211 | **1.59E-04** | 1466 | 0.013 | -0.05 | 0.75 |
| *RBM47* | PAV | apoC-III | 1.33E-06 | 4:40426074:C:T (rs35529250)*; | 0.001 | -1.020 | 0.31 | 2982 | 0.005 | 0.06 | 0.76 |
| | | | | 4:40432687:AGCGGCTGCGGCGGCTGCGGCC:A (rs564837143, in frame deletion) | 0.005 | -1.938 | **2.50E-06** | 2982 | 0.008 | 0.26 | 0.08 |
| | | | | 4:40438508:C:T (rs373211767*) | 0.002 | 0.817 | 0.25 | 2982 | 0.001 | 0.52 | 0.23 |

P_LIU: P-value for SKAT meta-analysis gene test. Variants: Variants within the gene that contributed to the SKAT meta-analysis (i.e. all PTVs or PAVs within the gene. Annotation includes chr:pos:REF:ALT alleles, rs number if known, and consequence. *indicates nonsynonymous missense mutation. MAF: Minor allele frequency for each variant; Effect and P-value: beta effect size estimate and p-value from score test meta-analysis. GWAS: N, Allele frequency (AF), Effect size and P-value for the variants in the non-overlapping GWAS data. P-values <0.05 (not corrected for multiple testing) are highlighted in bold.

VLDLPL XL: Phospholipid in extra large VLDL, VLDLPL XS: Phospholipid in extra small VLDL, apoC-III: Serum Apolipoprotein C-III, apoA1: Serum apolipoprotein A-I [mg/dl], CHOL CL: Total Cholesterol, LDL Friedewald: LDL cholesterol calculated with Friedewald equation. Non-HDLC: Non-HDL cholesterol.

**LIPC rs121912502 was imputed with low imputation quality, INFO 0.37

**Supplementary Table S6: Variant association with "Diseases of the circulatory system" phenotypes in the FinnGen GWAS data.**

| GENE: Pheno | VARs | Effect | rsID | MAFs | EFF | P-values | SIFT | PolyPhen | FinEnr | phenotype | beta | p-value | N |
|---|---|---|---|---|---|---|---|---|---|---|---|---|---|
| *DEFT1P/ DEFT1P2:* VLDLPL XL | 8:6989658:C:T | Splice donor | rs797006828 | 0.020 | 1.157 | **1.23E-06** | - | - | | | | | |
| *SBDS:* APOC3 | 7:66994210:A:G | Splice donor | rs113993993 | 0.005 | 1.865 | **5.86E-06** | - | - | 2.27 | Hypertension complicating pregnancy, childbirth, and the puerperium | 0.26 | 1.60E-02 | 6120 / 94241 |
| | | | | | | | | | | Varicose veins | -0.19 | 2.10E-02 | 13928 / 153951 |
| | | | | | | | | | | Vascular dementia | 0.7 | 2.50E-02 | 706 / 171075 |
| | | | | | | | | | | Sequelae of cerebrovascular disease | 0.31 | 3.00E-02 | 3520 / 165040 |
| | | | | | | | | | | Aortic aneurysm | -0.41 | 3.40E-02 | 1919 / 167843 |
| | | | | | | | | | | Secondary right heart disease | 0.94 | 3.60E-02 | 315 / 173597 |
| | | | | | | | | | | Diseases of veins, lymphatic vessels and lymph nodes, not elsewhere classified | -0.13 | 4.80E-02 | 22948 / 153951 |
| | 7:66994286:T:A | K/* | rs120074160 | 0.001 | 0.603 | 0.55 | - | - | | | | | |
| *LIPC:* APOA1 | 15:58541794:G:A | V/M | rs6078 | 0.032 | 0.171 | 0.20 | 1 | 0.01 | 1.79 | Venous thromboembolism | -0.11 | 4.00E-03 | 6913 / 169986 |
| | | | | | | | | | | DVT of lower extremities and pulmonary embolism | -0.12 | 5.50E-03 | 6019 / 170880 |
| | | | | | | | | | | Pulmonary heart disease, diseases of pulmonary circulation | -0.13 | 1.90E-02 | 3302 / 173597 |
| | | | | | | | | | | Aneurysms, operations, SAH | -0.18 | 2.40E-02 | 1582 / 165040 |
| | | | | | | | | | | Other diseases of arteries and capillaries | 0.21 | 2.90E-02 | 1046 / 167843 |
| | | | | | | | | | | Cardiomyopathy, other and unspecified | -0.22 | 3.00E-02 | 971 / 129908 |
| | | | | | | | | | | Pulmonary embolism | -0.12 | 3.50E-02 | 3016 / 173597 |
| | | | | | | | | | | Cerebral aneurysm, nonruptured | -0.25 | 3.70E-02 | 720 / 165040 |
| | | | | | | | | | | Nontraumatic intracranial haemmorrhage | -0.15 | 3.80E-02 | 2086 / 165040 |
| | | | | | | | | | | Other peripheral vascular diseases | -0.23 | 4.30E-02 | 799 / 167843 |
| | 15:58541843:C:T | P/L | rs562988299 | 0.001 | 0.877 | 0.38 | 0.31 | 0 | | | | | |
| | 15:58541944:G:A | A/T | rs201563586 | 0.001 | 1.794 | 0.07 | 0.01 | 0.97 | None | Non-rheumatic valve diseases | -0.48 | 2.10E-02 | 7440 / 129908 |
| | | | | | | | | | | Other other unspecified disorders of the circulatory system | 1.9 | 3.00E-02 | 340 / 175178 |
| | | | | | | | | | | Aortic aneurysm | -0.78 | 4.20E-02 | 1919 / 167843 |
| | 15:58548387:C:T | S/F | rs121912502 | 0.001 | 0.937 | 0.35 | 0 | 0.987 | 0.20 | Intracerebral haemmorrhage | 2.6 | 7.80E-03 | 1224 / 163533 |
| | | | | | | | | | | Transient ischemic attack | 0.96 | 9.20E-03 | 6729 / 164286 |
| | | | | | | | | | | Other intracranial haemorrhages | 6.1 | 1.20E-02 | 230 / 165040 |
| | | | | | | | | | | Cardiomyopathy, other and unspecified | 2.2 | 2.10E-02 | 971 / 129908 |
| | | | | | | | | | | Nontraumatic intracranial haemmorrhage | 1.3 | 3.10E-02 | 2086 / 165040 |
| | 15:58560880:A:C | F/L | rs3829462 | 0.045 | 0.001 | 0.99 | - | - | 0.96 | Other or ill-defined heart diseases | 0.33 | 1.3e-2 | 583 / 129908 |
| | | | | | | | | | | Other pulmonary heart/vessel disease | -0.37 | 2.9e-2 | 363 / 173597 |
| | | | | | | | | | | Secondary right heart disease | -0.39 | 3.5e-2 | 315 / 173597 |
| | | | | | | | | | | Phlebitis and thrombophlebitis (not including DVT) | -0.14 | 4.2e-2 | 2506 / 153951 |
| | 15:58563549:C:T | T/M | rs113298164 | 0.017 | 0.980 | **7.77E-08** | 0.02 | 0.996 | 4.41 | Secondary hypertension | 0.36 | 4.90E-02 | 982 / 133323 |

| | | | | | | | | | | | | |
|---|---|---|---|---|---|---|---|---|---|---|---|---|
| *GTF3C5:* | | | | | | | | | | | | |
| Non-HDLC | 9:133042147:C:T | H/Y | - | 0.001 | 0.596 | 0.55 | 0 | 0.998 | | | | |
| | 9:133043731:T:C | Start lost; M/T | rs189383196 | 0.004 | 0.311 | 0.38 | 0 | 0.932 | 84.2 | Non-ischemic cardiomyopathy | 0.33 | 2.80E-05 | 8300 / 143233 |
| | | | | | | | | | | Hypertension | 0.16 | 6.70E-04 | 43545 / 133323 |
| | | | | | | | | | | Hypertensive diseases | 0.15 | 7.50E-04 | 43576 / 133323 |
| | | | | | | | | | | Cardiovascular diseases | 0.12 | 1.90E-03 | 86957 / 89942 |
| | | | | | | | | | | Statin medication | 0.14 | 2.10E-03 | 53518 / 123381 |
| | | | | | | | | | | DVT of lower extremities | 0.35 | 2.20E-03 | 3592 / 153951 |
| | | | | | | | | | | All-cause Heart Failure | 0.17 | 4.50E-03 | 17387 / 159058 |
| | | | | | | | | | | Secondary right heart disease | 1 | 4.80E-03 | 315 / 173597 |
| | | | | | | | | | | Venous thromboembolism | 0.24 | 5.30E-03 | 6913 / 169986 |
| | | | | | | | | | | Heart failure, not strict | 0.16 | 6.70E-03 | 17613 / 159286 |
| | | | | | | | | | | Hypertension, essential | 0.13 | 7.20E-03 | 33229 / 133323 |
| | | | | | | | | | | Phlebitis and thrombophlebitis (not including DVT) | 0.37 | 7.90E-03 | 2506 / 153951 |
| | | | | | | | | | | Other heart diseases | 0.1 | 1.00E-02 | 46991 / 129908 |
| | | | | | | | | | | Peripheral artery operations in Hilmo | 0.45 | 1.70E-02 | 1254 / 167843 |
| | | | | | | | | | | Conduction disorders | 0.29 | 1.80E-02 | 3271 / 129908 |
| | | | | | | | | | | Other pulmonary heart/vessel disease | 0.79 | 1.90E-02 | 363 / 173597 |
| | | | | | | | | | | Diseases of arteries, arterioles and capillaries (FINNGEN) | 0.22 | 2.80E-02 | 6988 / 97214 |
| | | | | | | | | | | Diseases of veins, lymphatic vessels and lymph nodes, not elsewhere classified | 0.11 | 2.90E-02 | 22948 / 153951 |
| | | | | | | | | | | Atrial fibrillation and flutter | 0.17 | 3.00E-02 | 17325 / 97214 |
| | | | | | | | | | | Diseases of arteries, arterioles and capillaries | 0.15 | 4.70E-02 | 9056 / 167843 |
| | 9:133043838:C:A | Q/K | rs150056568 | 0.001 | 0.017 | 0.98 | 0.08 | 0.755 | 2.30 | Non-ischemic cardiomyopathy | 0.46 | 7.80E-03 | 8300 / 143233 |
| | | | | | | | | | | Nonischemic cardiomyopathy | 1.5 | 1.20E-02 | 653 / 150661 |
| | | | | | | | | | | Other diseases of arteries and capillaries | -1 | 2.40E-02 | 1046 / 167843 |
| | | | | | | | | | | Transient ischemic attack | -0.42 | 2.60E-02 | 6729 / 164286 |
| | | | | | | | | | | Nonspesific lymphadenitis | 1.1 | 2.60E-02 | 806 / 153951 |
| | | | | | | | | | | Ischaemic Stroke, excluding all haemorrhages | -0.35 | 4.70E-02 | 8046 / 164286 |
| | | | | | | | | | | Cardiomyopathy | 0.61 | 4.80E-02 | 2342 / 129908 |
| | 9:133054459:A:G | Y/C | rs369889499 | 0.001 | -0.392 | 0.58 | 0 | 0.993 | None | Angina pectoris | 1 | 9.20E-05 | 14712 / 152621 |
| | | | | | | | | | | Ischaemic heart disease, wide definition | 0.71 | 6.10E-04 | 25366 / 151533 |
| | | | | | | | | | | Unstable angina pectoris | 1.2 | 2.40E-03 | 5774 / 160163 |
| | | | | | | | | | | Ischemic heart diseases | 0.62 | 3.10E-03 | 24278 / 152621 |
| | | | | | | | | | | Cardiomyopathies, Primary/intrinsic | 1.7 | 3.40E-03 | 1678 / 129908 |
| | | | | | | | | | | Other diseases of pericardium | 5.1 | 4.50E-03 | 307 / 129908 |
| | | | | | | | | | | Cardiomyopathy, Hypertrophic obstructive | 4.7 | 7.00E-03 | 251 / 129908 |
| | | | | | | | | | | Coronary revascularization (ANGIO or CABG) | 0.79 | 1.20E-02 | 9971 / 152621 |
| | | | | | | | | | | Myocarditis | 2.7 | 1.30E-02 | 626 / 97214 |
| | | | | | | | | | | Other CVD (FINNGEN) | 2 | 1.40E-02 | 1398 / 97214 |

| | | | | | | | | | | | | | |
|---|---|---|---|---|---|---|---|---|---|---|---|---|---|
| | | | | | | | | | | Other disorders of veins | -0.88 | 2.10E-02 | 4588 / 153951 |
| | | | | | | | | | | Valvular operations | 0.39 | 2.30E-02 | 26015 / 129908 |
| | | | | | | | | | | Valvular heart disease including rheumatic fever | 0.38 | 2.30E-02 | 27684 / 129908 |
| | | | | | | | | | | Cardiomyopathy | 1 | 3.00E-02 | 2342 / 129908 |
| | | | | | | | | | | Left bundle-branch block | 2.4 | 3.20E-02 | 582 / 129908 |
| | | | | | | | | | | Cardiovascular diseases | 0.29 | 3.60E-02 | 86957 / 89942 |
| | | | | | | | | | | Coronary atherosclerosis | 0.51 | 3.70E-02 | 18295 / 152621 |
| | | | | | | | | | | Hypertrophic cardiomyopathy | 2.2 | 3.80E-02 | 432 / 176467 |
| | | | | | | | | | | Endocarditis | 2.7 | 4.70E-02 | 312 / 129908 |
| | | | | | | | | | | Heart failure,strict | 0.54 | 4.80E-02 | 9576 / 159286 |
| | | | | | | | | | | Hypertension | 0.33 | 4.90E-02 | 43545 / 133323 |
| | 9:133054733:A:G | H/R | rs1302997790 | 0.001 | 0.840 | 0.40 | 0.5 | 0.011 | | | | | |
| | 9:133054787:C:T | A/V | rs202207045 | 0.007 | -1.382 | **7.26E-07** | 0.24 | 0.03 | None | Vascular dementia | -0.76 | 2.10E-02 | 706 / 171075 |
| | 9:133056848:G:A | D/N | rs637435 | 0.001 | -0.596 | 0.55 | 0.04 | 0.833 | | | | | |
| | 9:133057819:T:C | F/L | rs111893665 | 0.001 | -0.120 | 0.90 | 0.27 | 0.914 | 0.152* | Other peripheral vascular diseases | 2.9 | 1.00E-02 | 799 / 167843 |
| | | | | | | | | | | Paroxysmal tachycardia | -1.1 | 2.10E-02 | 3667 / 97214 |
| *TRMT5:* VLDLPL_XS | 14:60975180:G:A | P/S | rs147405788 | 0.001 | -1.575 | 0.03 | 0 | 0.88 | 6.24 | Type 2 diabetes with peripheral circulatory complications | -1.1 | 3.40E-02 | 820 / 148190 |
| | | | | | | | | | | Cardiomyopathy, Hypertrophic obstructive | 1.7 | 3.60E-02 | 251 / 129908 |
| | 14:60975182:T:C | D/G | - | 0.001 | -1.386 | 0.17 | 0.1 | 0.001 | | | | | |
| | 14:60975636:G:A | A/V | rs45604437 | 0.007 | -1.094 | **5.76E-04** | 0.16 | 0 | 2.73 | Other peripheral vascular diseases | 1.1 | 4.90E-03 | 799 / 167843 |
| | | | | | | | | | | Phlebitis and thrombophlebitis (not including DVT) | 0.5 | 1.90E-02 | 2506 / 153951 |
| | | | | | | | | | | Sequelae of cerebrovascular disease | -0.44 | 2.10E-02 | 3520 / 165040 |
| | | | | | | | | | | Hypertensive Renal Disease | 1.2 | 2.60E-02 | 373 / 133323 |
| | | | | | | | | | | Stroke, excluding SAH | -0.25 | 4.60E-02 | 8877 / 163535 |
| | | | | | | | | | | Stroke, including SAH | -0.23 | 4.90E-02 | 9592 / 162861 |
| | 14:60979242:T:C | K/R | rs138139551 | 0.002 | -0.599 | 0.30 | 0.2 | 0.253 | None | Other noninfective disordersof lymphatic vessels and lymph nodes | 2.8 | 3.00E-03 | 301 / 153951 |
| | | | | | | | | | | Left bundle-branch block | 1.6 | 1.30E-02 | 582 / 129908 |
| | | | | | | | | | | Occlusion and stenosis of arteries, not leading to stroke | 2.7 | 2.50E-02 | 147 / 165040 |
| | | | | | | | | | | DVT of lower extremities | -0.58 | 2.70E-02 | 3592 / 153951 |
| | | | | | | | | | | Status post-ami | 0.96 | 2.70E-02 | 1141 / 152621 |
| | | | | | | | | | | Hypertensive Renal Disease | 1.8 | 3.40E-02 | 373 / 133323 |
| | | | | | | | | | | Conduction disorders | 0.57 | 3.70E-02 | 3271 / 129908 |
| | 14:60979257:T:C | H/R | rs191207997 | 0.001 | -0.713 | 0.48 | 0.05 | 0.27 | 3.12 | Hypertensive Heart Disease | 0.95 | 1.40E-02 | 3252 / 133323 |
| | | | | | | | | | | Non-rheumatic valve diseases | 0.62 | 1.50E-02 | 7440 / 129908 |
| | 14:60979303:C:G | G/R | rs1285433520 | 0.001 | 0.400 | 0.69 | 0 | 0.588 | | | | | |
| | 14:60979401:G:A | P/L | rs746471581 | 0.001 | 1.362 | 0.17 | 0.02 | 0.031 | | | | | |

| | | | | | | | | | | | | | |
|---|---|---|---|---|---|---|---|---|---|---|---|---|---|
| | 14:60979429:T:A | S/C | rs115400838 | 0.007 | -1.211 | **1.59E-04** | 0.03 | 0.375 | 6.99 | Stroke, excluding SAH | 0.24 | 1.90E-04 | 8877 / 163535 |
| | | | | | | | | | | Stroke, including SAH | 0.22 | 3.70E-04 | 9592 / 162861 |
| | | | | | | | | | | Sequelae of cerebrovascular disease | 0.35 | 5.40E-04 | 3520 / 165040 |
| | | | | | | | | | | Ischaemic Stroke, excluding all haemorrhages | 0.23 | 6.00E-04 | 8046 / 164286 |
| | | | | | | | | | | Cerebrovascular diseases | 0.18 | 1.60E-03 | 11859 / 165040 |
| | | | | | | | | | | Cerebrovascular diseases (FINNGEN) | 0.17 | 1.40E-02 | 10367 / 97214 |
| | | | | | | | | | | Arterial embolism and thrombosis | 0.58 | 1.50E-02 | 580 / 167843 |
| | | | | | | | | | | Rheumatic fever incl heart disease | 0.56 | 3.30E-02 | 462 / 176437 |
| | | | | | | | | | | Valvular operations | 0.087 | 3.50E-02 | 26015 / 129908 |
| *RBM47:* APOC3 | 4:40426074:C:T | G/R | rs35529250 | 0.001 | -1.020 | 0.31 | 0 | 0.905 | 0.81 | Cardiomyopathy, Hypertrophic obstructive | 2.3 | 2.10E-03 | 251 / 129908 |
| | | | | | | | | | | Hypertrophic cardiomyopathy | 1.4 | 7.30E-03 | 432 / 176467 |
| | | | | | | | | | | Hypertension, essential | -0.18 | 1.60E-02 | 33229 / 133323 |
| | | | | | | | | | | Hypertensive diseases | -0.17 | 1.90E-02 | 43576 / 133323 |
| | | | | | | | | | | Hypertension | -0.17 | 1.90E-02 | 43545 / 133323 |
| | | | | | | | | | | Phlebitis and thrombophlebitis (not including DVT) | -0.45 | 2.30E-02 | 2506 / 153951 |
| | | | | | | | | | | Diseases of veins, lymphatic vessels and lymph nodes, not elsewhere classified | -0.17 | 3.50E-02 | 22948 / 153951 |
| | | | | | | | | | | Stroke, excluding SAH | -0.24 | 3.70E-02 | 8877 / 163535 |
| | | | | | | | | | | Ischaemic Stroke, excluding all haemorrhages | -0.24 | 4.30E-02 | 8046 / 164286 |
| | 4:40432687:AGC GGCTGCGGCGG CTGCGGCC:A | AAAAAAAA/A | rs564837143 | 0.005 | -1.938 | **2.50E-06** | - | - | | Secondary hypertension | -0.65 | 1.10E-02 | 982 / 133323 |
| | | | | | | | | | | Other peripheral vascular diseases | -0.58 | 3.50E-02 | 799 / 167843 |
| | 4:40438508:C:T | R/H | rs373211767 | 0.002 | 0.817 | 0.25 | 0.03 | 0.891 | None | Vascular syndromes of brain in cerebrovascular disorders | 3.4 | 6.90E-03 | 458 / 170687 |
| | | | | | | | | | | Angina pectoris | 0.58 | 1.20E-02 | 14712 / 152621 |
| | | | | | | | | | | Aortic aneurysm | 1.1 | 2.80E-02 | 1919 / 167843 |
| | | | | | | | | | | Status post-ami | 1.4 | 2.80E-02 | 1141 / 152621 |
| | | | | | | | | | | Other specified cerebrovascular diseases, other cerebrovascular disorders in diseases classified elsewhere | 1.9 | 3.10E-02 | 729 / 165040 |
| | | | | | | | | | | Other aneurysm | 2.1 | 4.00E-02 | 444 / 167843 |

**Supplemental Table S7. Significant associations (WES/WGS SKAT meta-analysis p<0.05/19 genes=0.0026) for PTVs and PAVs in known lipid genes.**

| GENE | | Variants | Pheno | MAFs | SINGLEVAR EFFECTs | SINGLEVAR PVALUEs | P-VALUE | Classification |
|---|---|---|---|---|---|---|---|---|
| *APOB* | PTV | 2:21007222:AT:A (rs1232943044, frameshift)<br>2:21011037:TGA:T (rs1407451220, frameshift) | LDL Friedewald | 0.0006<br>0.001 | -0.46<br>-2.4 | 0.64<br>0.001 | 0.001 | Likely Pathogenic (PVS1, PM2)<br>Likely Pathogenic (PVS1, PM2) |
| *APOB* | PTV | 2:21007222:AT:A (rs1232943044, frameshift)<br>2:21011037:TGA:T (rs1407451220, frameshift) | Non-HDLC | 0.0005<br>0.001 | -0.64<br>-2.51 | 0.52<br>0.0004 | 0.0005 | Likely Pathogenic (PVS1, PM2)<br>Likely Pathogenic (PVS1, PM2) |
| *APOB* | PTV | 2:21007222:AT:A (rs1232943044, frameshift)<br>2:21011037:TGA:T (rs1407451220, frameshift) | APOB | 0.0005<br>0.001 | -1.63<br>-2.36 | 0.10<br>0.001 | 0.0006 | Likely Pathogenic (PVS1, PM2)<br>Likely Pathogenic (PVS1, PM2) |
| *APOB* | PTV | 2:21007222:AT:A (rs1232943044, frameshift)<br>2:21011037:TGA:T (rs1407451220, frameshift) | VLDLTG S | 0.0007<br>0.001 | -1.19<br>-2.32 | 0.23<br>0.001 | 0.001 | Likely Pathogenic (PVS1, PM2)<br>Likely Pathogenic (PVS1, PM2) |
| *PCSK9* | PAV | 1:55039974:G:T(rs11591147*)<br>1:55039995:C:T(rs11583680*)<br>1:55052701:C:T(rs148195424*) | LDL Friedewald | 0.02<br>0.05<br>0.001 | -0.56<br>-0.10<br>-1.14 | 0.001<br>0.37<br>0.11 | 0.001 | Benign (BP6, BS1, BS2, BP1)<br>Benign (BA1, BP6, BP4, BP1)<br>Benign (BS1, BS2, PP3, BP6, BP1) |
| *PCSK9* | PAV | 1:55039974:G:T(rs11591147*)<br>1:55039995:C:T(rs11583680*)<br>1:55052701:C:T(rs148195424*) | Non-HDLC | 0.02<br>0.05<br>0.001 | -0.61<br>-0.07<br>-0.93 | 0.0004<br>0.55<br>0.19 | 0.0005 | Benign (BP6, BS1, BS2, BP1)<br>Benign (BA1, BP6, BP4, BP1)<br>Benign (BS1, BS2, PP3, BP6, BP1) |
| *PCSK9* | PAV | 1:55039974:G:T(rs11591147*)<br>1:55039995:C:T(rs11583680*)<br>1:55052701:C:T(rs148195424*) | Total CHOL | 0.02<br>0.05<br>0.001 | -0.64<br>-0.06<br>-0.44 | 0.0002<br>0.63<br>0.54 | 0.0003 | Benign (BP6, BS1, BS2, BP1)<br>Benign (BA1, BP6, BP4, BP1)<br>Benign (BS1, BS2, PP3, BP6, BP1) |
| *PCSK9* | PAV | 1:55039974:G:T(rs11591147*)<br>1:55039995:C:T(rs11583680*) | Isoleucine | 0.02<br>0.04 | -0.75<br>0.15 | 0.0003<br>0.25 | 0.0003 | Benign (BS2, BP6, BP4, BP1, BS1)<br>Benign (BP4, BA1, BP6, BP1) |
| *LIPC* | PAV | 15:58541794:G:A (rs6078*)<br>15:58541843:C:T(rs562988299*)<br>15:58541944:G:A(rs201563586*)<br>15:58548387:C:T(rs121912502*)<br>15:58560880:A:C*<br>15:58563549:C:T(rs113298164*) | APOA1 | 0.03<br>0.0006<br>0.006<br>0.0006<br>0.05<br>0.02 | 0.17<br>0.88<br>1.80<br>0.94<br>0.001<br>0.98 | 0.20<br>0.38<br>0.07<br>0.35<br>0.99<br>7.78E-08 | 1.48E-07 | Benign (BA1, BP4, BP1, BP6)<br>Likely Benign (PM2, BP1, BP4)<br>VUS (PM2, PP3, BP1)<br>VUS (PP3, PM2, PP5, BP1)<br>-<br>Benign (BS1, BS2, BP1, PP3, PP5) |
| *LIPC* | PAV | 15:58541794:G:A (rs6078*)<br>15:58541843:C:T(rs562988299*)<br>15:58541944:G:A(rs201563586*)<br>15:58548387:C:T(rs121912502*)<br>15:58560880:A:C*<br>15:58563549:C:T(rs113298164*) | HDL CHOL | 0.03<br>0.0005<br>0.0005<br>0.0005<br>0.05<br>0.02 | 0.17<br>-0.51<br>0.09<br>0.22<br>0.04<br>0.61 | 0.21<br>0.61<br>0.93<br>0.82<br>0.75<br>0.001 | 0.002 | Benign (BA1, BP4, BP1, BP6)<br>Likely Benign (PM2, BP1, BP4)<br>VUS (PM2, PP3, BP1)<br>VUS (PP3, PM2, PP5, BP1)<br>-<br>Benign (BS1, BS2, BP1, PP3, PP5) |
| *LIPC* | PAV | 15:58541794:G:A (rs6078*)<br>15:58541843:C:T(rs562988299*)<br>15:58541944:G:A(rs201563586*)<br>15:58548387:C:T(rs121912502*)<br>15:58560880:A:C*<br>15:58563549:C:T(rs113298164*) | HDLCE XL | 0.03<br>0.0007<br>0.0007<br>0.0007<br>0.05<br>0.02 | 0.19<br>0.76<br>1.71<br>2.07<br>0.11<br>0.75 | 0.20<br>0.45<br>0.09<br>0.04<br>0.39<br>7.22E-05 | 0.0001 | Benign (BA1, BP4, BP1, BP6)<br>Likely Benign (PM2, BP1, BP4)<br>VUS (PM2, PP3, BP1)<br>VUS (PP3, PM2, PP5, BP1)<br>-<br>Benign (BS1, BS2, BP1, PP3, PP5) |

| | | | | | | | | |
|---|---|---|---|---|---|---|---|---|
| *LIPC* | PAV | 15:58541794:G:A (rs6078*)<br>15:58541843:C:T(rs562988299*)<br>15:58541944:G:A(rs201563586*)<br>15:58548387:C:T(rs121912502*)<br>15:58560880:A:C*<br>15:58563549:C:T(rs113298164*) | HDLC XL | 0.03<br>0.0007<br>0.0007<br>0.0007<br>0.05<br>0.02 | 0.16<br>0.77<br>1.43<br>1.78<br>0.07<br>0.80 | 0.30<br>0.44<br>0.15<br>0.07<br>0.61<br>2.65E-05 | 8.12E-05 | Benign (BA1, BP4, BP1, BP6)<br>Likely Benign (PM2, BP1, BP4)<br>VUS (PM2, PP3, BP1)<br>VUS (PP3, PM2, PP5, BP1)<br>Benign (BS1, BS2, BP1, PP3, PP5) |
| *LIPC* | PAV | 15:58541794:G:A (rs6078*)<br>15:58541843:C:T(rs562988299*)<br>15:58541944:G:A(rs201563586*)<br>15:58548387:C:T(rs121912502*)<br>15:58560880:A:C*<br>15:58563549:C:T(rs113298164*) | HDLFC XL | 0.03<br>0.0007<br>0.0007<br>0.0007<br>0.05<br>0.02 | 0.04<br>0.47<br>0.57<br>0.87<br>-0.02<br>0.68 | 0.79<br>0.64<br>0.57<br>0.38<br>0.88<br>0.0003 | 0.002 | Benign (BA1, BP4, BP1, BP6)<br>Likely Benign (PM2, BP1, BP4)<br>VUS (PM2, PP3, BP1)<br>VUS (PP3, PM2, PP5, BP1)<br>-<br>Benign (BS1, BS2, BP1, PP3, PP5) |
| *LIPC* | PAV | 15:58541794:G:A (rs6078*)<br>15:58541843:C:T(rs562988299*)<br>15:58541944:G:A(rs201563586*)<br>15:58548387:C:T(rs121912502*)<br>15:58560880:A:C*<br>15:58563549:C:T(rs113298164*) | HDLTG XL | 0.03<br>0.0007<br>0.0007<br>0.0007<br>0.05<br>0.02 | 0.39<br>1.13<br>1.15<br>0.90<br>0.03<br>0.52 | 0.009<br>0.26<br>0.25<br>0.37<br>0.85<br>0.006 | 0.001 | Benign (BA1, BP4, BP1, BP6)<br>Likely Benign (PM2, BP1, BP4)<br>VUS (PM2, PP3, BP1)<br>VUS (PP3, PM2, PP5, BP1)<br>-<br>Benign (BS1, BS2, BP1, PP3, PP5) |
| *LIPC* | PAV | 15:58541794:G:A (rs6078*)<br>15:58541843:C:T(rs562988299*)<br>15:58541944:G:A(rs201563586*)<br>15:58548387:C:T(rs121912502*)<br>15:58560880:A:C*<br>15:58563549:C:T(rs113298164*) | HDL XL | 0.03<br>0.0007<br>0.0007<br>0.0007<br>0.05<br>0.02 | 0.15<br>0.65<br>0.70<br>1.19<br>0.01<br>0.79 | 0.30<br>0.52<br>0.48<br>0.23<br>0.92<br>2.74E-05 | 0.0001 | Benign (BA1, BP4, BP1, BP6)<br>Likely Benign (PM2, BP1, BP4)<br>VUS (PM2, PP3, BP1)<br>VUS (PP3, PM2, PP5, BP1)<br>-<br>Benign (BS1, BS2, BP1, PP3, PP5) |
| *LIPC* | PAV | 15:58541794:G:A (rs6078*)<br>15:58541843:C:T(rs562988299*)<br>15:58541944:G:A(rs201563586*)<br>15:58548387:C:T(rs121912502*)<br>15:58560880:A:C*<br>15:58563549:C:T(rs113298164*) | IDLTG | 0.03<br>0.0007<br>0.0007<br>0.0007<br>0.05<br>0.02 | 0.33<br>1.07<br>1.59<br>1.21<br>0.06<br>0.63 | 0.03<br>0.28<br>0.11<br>0.23<br>0.65<br>0.0009 | 0.0004 | Benign (BA1, BP4, BP1, BP6)<br>Likely Benign (PM2, BP1, BP4)<br>VUS (PM2, PP3, BP1)<br>VUS (PP3, PM2, PP5, BP1)<br>-<br>Benign (BS1, BS2, BP1, PP3, PP5) |
| *CETP* | PAV | 16:56975116:T:C*<br>16:56981179:G:C(rs5880*)<br>16:56983407:G:A(rs1800777*) | APOA1 | 0.0006<br>0.03<br>0.01 | 0.05<br>-0.58<br>-0.57 | 0.99<br>1.33E-05<br>0.006 | 5.62E-05 | Likely Benign (PM2, BP4, BP1)<br>Benign (BA1, BP6, BP1, BP4)<br>Benign (BA1, BP1, BP4, BP6) |
| *CETP* | PAV | 16:56975116:T:C*<br>16:56981179:G:C(rs5880*)<br>16:56983407:G:A(rs1800777*) | HDL2C | 0.0006<br>0.03<br>0.01 | -0.26<br>-0.48<br>-0.34 | 0.80<br>0.0003<br>0.12 | 0.002 | Likely Benign (PM2, BP4, BP1)<br>Benign (BA1, BP6, BP1, BP4)<br>Benign (BA1, BP1, BP4, BP6) |
| *CETP* | PAV | 16:56975116:T:C*<br>16:56981179:G:C(rs5880*)<br>16:56983407:G:A(rs1800777*) | HDL CHOL | 0.0005<br>0.04,<br>0.01 | -0.20<br>-0.52<br>-0.45 | 0.84<br>8.43E-05<br>0.03 | 0.0005 | Likely Benign (PM2, BP4, BP1)<br>Benign (BA1, BP6, BP1, BP4)<br>Benign (BA1, BP1, BP4, BP6) |
| *CETP* | PAV | 16:56975116:T:C*<br>16:56981179:G:C(rs5880*)<br>16:56983407:G:A(rs1800777*) | HDLCE M | 0.0007<br>0.03<br>0.008 | 0.94<br>-0.52<br>-0.78 | 0.35<br>0.001<br>0.008 | 0.0007 | Likely Benign (PM2, BP4, BP1)<br>Benign (BA1, BP6, BP1, BP4)<br>Benign (BA1, BP1, BP4, BP6) |
| *CETP* | PAV | 16:56975116:T:C*<br>16:56981179:G:C(rs5880*)<br>16:56983407:G:A(rs1800777*) | HDLC M | 0.0007<br>0.03<br>0.008 | 1.02<br>-0.57<br>-0.82 | 0.31<br>0.0004<br>0.005 | 0.0002 | Likely Benign (PM2, BP4, BP1)<br>Benign (BA1, BP6, BP1, BP4)<br>Benign (BA1, BP1, BP4, BP6) |
| *CETP* | PAV | 16:56975116:T:C*<br>16:56981179:G:C(rs5880*)<br>16:56983407:G:A(rs1800777*) | HDLC | 0.0007<br>0.03<br>0.008 | -0.02<br>-0.61<br>-1.02 | 0.99<br>0.0002<br>0.0005 | 3.06E-05 | Likely Benign (PM2, BP4, BP1)<br>Benign (BA1, BP6, BP1, BP4)<br>Benign (BA1, BP1, BP4, BP6) |
| *CETP* | PAV | 16:56975116:T:C*<br>16:56981179:G:C(rs5880*)<br>16:56983407:G:A(rs1800777*) | HDLFC M | 0.0007<br>0.03<br>0.008 | 0.92<br>-0.48<br>-0.72 | 0.36<br>0.003<br>0.01 | 0.002 | Likely Benign (PM2, BP4, BP1)<br>Benign (BA1, BP6, BP1, BP4)<br>Benign (BA1, BP1, BP4, BP6) |
| *CETP* | PAV | 16:56975116:T:C*<br>16:56981179:G:C(rs5880*)<br>16:56983407:G:A(rs1800777*) | HDL M | 0.0007<br>0.03<br>0.008 | 1.02<br>-0.57<br>-0.82 | 0.31<br>0.0004<br>0.005 | 0.0002 | Likely Benign (PM2, BP4, BP1)<br>Benign (BA1, BP6, BP1, BP4)<br>Benign (BA1, BP1, BP4, BP6) |

| | | | | | | | | |
|---|---|---|---|---|---|---|---|---|
| *CETP* | PAV | 16:56975116:T:C*<br>16:56981179:G:C(rs5880*)<br>16:56983407:G:A(rs1800777*) | HDLPL M | 0.0007<br>0.03<br>0.008 | 0.15<br>-0.51<br>-0.95 | 0.88<br>0.002<br>0.001 | 0.0003 | Likely Benign (PM2, BP4, BP1)<br>Benign (BA1, BP6, BP1, BP4)<br>Benign (BA1, BP1, BP4, BP6) |
| *APOE* | PAV | 19:44907807:G:A(rs201672011*)<br>19:44907853:T:C(rs769452*)<br>19:44908822:C:T (rs7412*) | APOB | 0.0006<br>0.009<br>0.04 | 0.11<br>0.31<br>-0.49 | 0.91<br>0.21<br>0.0001 | 0.0003 | VUS (PM2, PP2, PP5, BP4)<br>Likely Benign (PP2, BP4, BS2)<br>VUS (PM1, PP2, PP3, BA1, PP5) |
| *APOE* | PAV | 19:44907853:T:C (rs201672011*)<br>19:44908822:C:T(rs7412*) | HDLD | 0.005<br>0.03 | -0.37<br>1.16 | 0.60<br>0.0005 | 0.0007 | Likely Benign (PP2, BS2, BP4)<br>VUS (PM2, PP2, PP3, PP5, BA1) |
| *APOE* | PAV | 19:44907807:G:A(rs201672011*)<br>19:44907853:T:C(rs769452*)<br>19:44908822:C:T (rs7412*) | LDLCE L | 0.0007<br>0.009<br>0.03 | -2.43<br>0.26<br>-0.57 | 0.01<br>0.35<br>0.00012 | 0.0002 | VUS (PM2, PP2, PP5, BP4),<br>Likely Benign (PP2, BP4, BS2)<br>VUS (PM1, PP2, PP3, BA1, PP5) |
| *APOE* | PAV | 19:44907807:G:A(rs201672011*)<br>19:44907853:T:C(rs769452*)<br>19:44908822:C:T (rs7412*) | LDLCE M | 0.0007<br>0.009<br>0.03 | -0.61<br>0.15<br>-0.53 | 0.54<br>0.59<br>0.0005 | 0.001 | VUS (PM2, PP2, PP5, BP4)<br>Likely Benign (PP2, BP4, BS2)<br>VUS (PM1, PP2, PP3, BA1, PP5) |
| *APOE* | PAV | 19:44907807:G:A(rs201672011*)<br>19:44907853:T:C(rs769452*)<br>19:44908822:C:T (rs7412*) | LDLC L | 0.0007<br>0.009<br>0.03 | -2.44<br>0.30<br>-0.58 | 0.01<br>0.29<br>0.0001 | 0.0002 | VUS (PM2, PP2, PP5, BP4)<br>Likely Benign (PP2, BP4, BS2)<br>VUS (PM1, PP2, PP3, BA1, PP5) |
| *APOE* | PAV | 19:44907807:G:A(rs201672011*)<br>19:44907853:T:C(rs769452*)<br>19:44908822:C:T (rs7412*) | LDLC M | 0.0007<br>0.009<br>0.03 | -1.08<br>0.25<br>-0.58 | 0.28<br>0.38<br>0.0001 | 0.0003 | VUS (PM2, PP2, PP5, BP4)<br>Likely Benign (PP2, BP4, BS2)<br>VUS (PM1, PP2, PP3, BA1, PP5) |
| *APOE* | PAV | 19:44907807:G:A(rs201672011*)<br>19:44907853:T:C(rs769452*)<br>19:44908822:C:T (rs7412*) | LDLC | 0.0007<br>0.009<br>0.03 | -2.01<br>0.26<br>-0.57 | 0.04<br>0.35<br>0.0002 | 0.0003 | VUS (PM2, PP2, PP5, BP4)<br>Likely Benign (PP2, BP4, BS2)<br>VUS (PM1, PP2, PP3, BA1, PP5) |

PTV= Protein truncating variant. PAV= Protein altering variant. SINGLEVAR EFFECTs= Effect sizes of single variants. P-value=P-value for SKAT meta-analysis gene test. *indicates nonsynonymous missense mutation. Classification = Variant classification according to ACMG guidelines (*Richards et al. 2015*), using automated classification provided by VarSome (*Kopanos et al, 2019*):

PVS1: Pathogenic, Very Strong.
PS1-3: Pathogenic, Strong
PM1-5: Pathogenic, Moderate.
PP2-5: Pathogenic, Supporting.
BA1: Benign, Stand Alone
BP1-7: Benign, Supporting
BS1-3: Benign, Strong

The studied genes include genes causing

- hypercholesteremia (*PCSK9* (MIM:607786), *LDLR* (MIM:606945), *APOB* (MIM:107730), *APOE* (MIM:107741), *LPA* (MIM:152200), *LDLRAP1* (MIM:605747), *LIPA* (MIM:613497), and *CYP7A1* (MIM:118455),
- monogenic hypertriglyceridemia (*LPL (MIM:609708)*, *APOC2* (MIM:608083), *GPIHBP1* (MIM: 612757), *APOA5* (MIM:606368) and *LMF1* (MIM:611761)),
- genetic disorders of HDL metabolism (*ABCA1* (MIM:600046), *LCAT* (MIM:606967), *APOA1* (MIM:107680), *ANGPTL3* (MIM:604774), *CETP* (MIM:118470), *LIPC* (MIM:151670), and *LIPG* (MIM: 603684))

as described in *Musunuru et al., 2015*.

**Supplementary Figure S1: WES+WGS SKAT meta-analysis results, Manhattan and QQ-plots.** A: Total cholesterol. B: Apo-A1. C: Phospholipids in extra large VLDL particles.

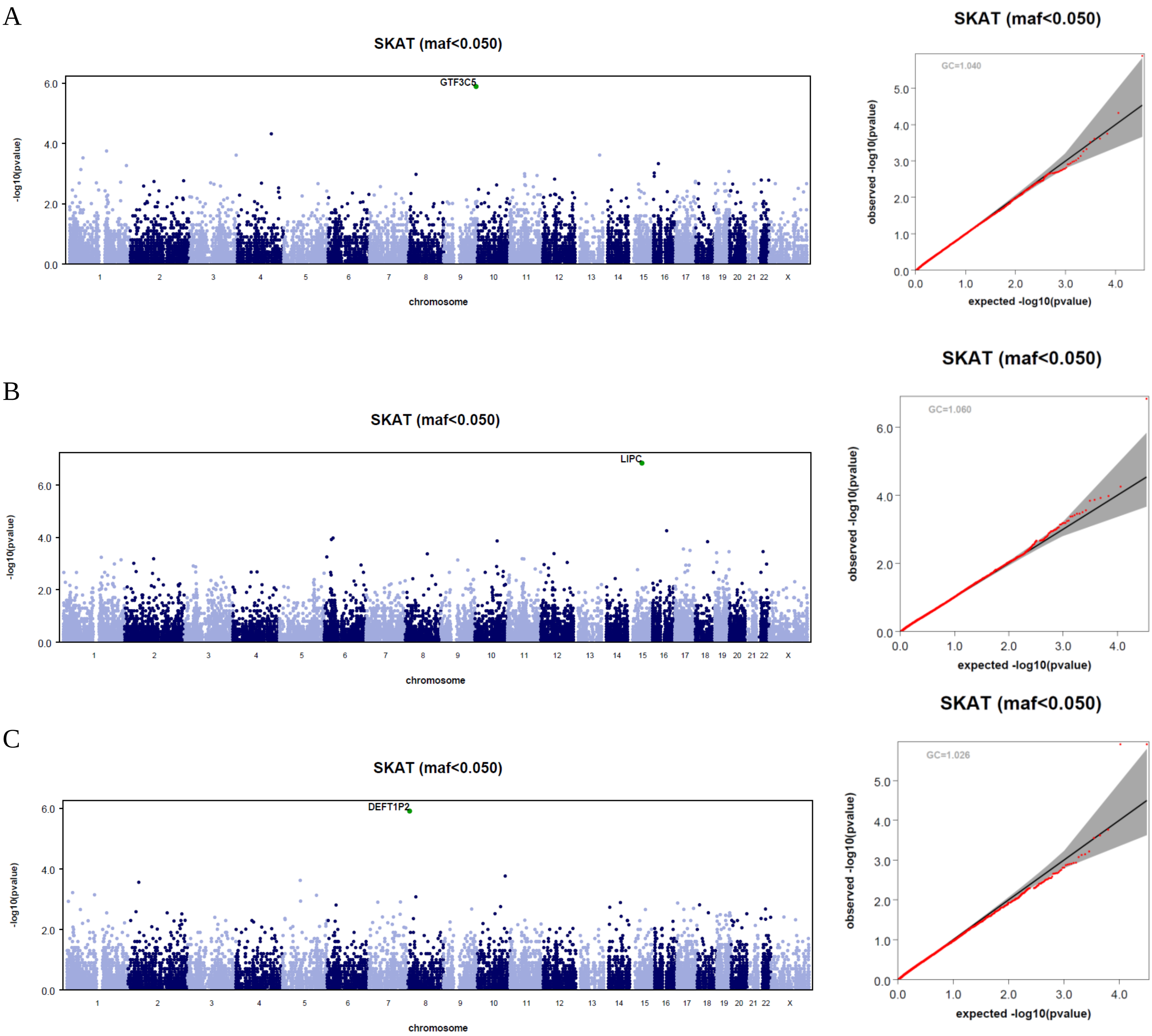

**Supplementary Figure S2: Apolipoprotein B (apoB) concentration in the three *APOB* rs1407451223 (N=2) or rs1232943044 (N=1) frameshift carriers (group 1; 44 – 50 mg/dl) vs. in the other participants (group 0; median 88 mg/dl, interquartile range 72 – 104 mg/dl).**

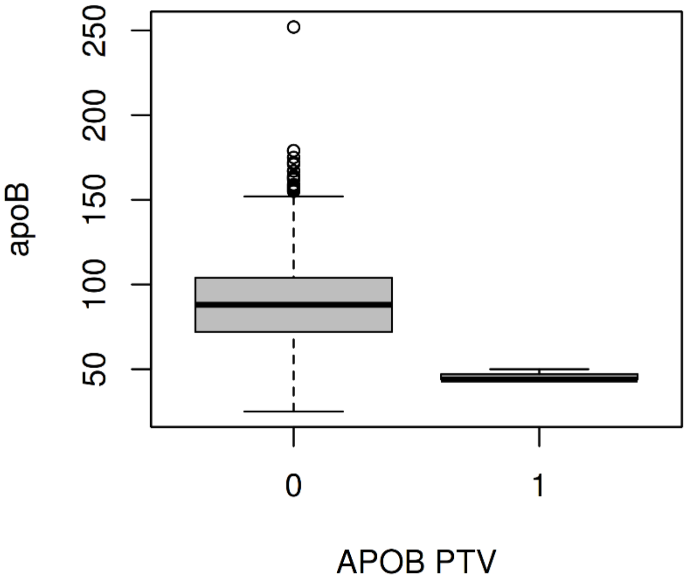

**STrengthening the REporting of Genetic Association studies (STREGA) reporting recommendations, extended from STROBE Statement**

| Item | Item no | STROBE Guideline | Extension for Genetic Association Studies (STREGA) | Page no |
|---|---|---|---|---|
| **Title and Abstract** | 1 | (a) Indicate the study's design with a commonly used term in the title or the abstract. | | 2 |
| | | (b) Provide in the abstract an informative and balanced summary of what was done and what was found. | | 2 |
| **Introduction** | | | | |
| *Background rationale* | 2 | Explain the scientific background and rationale for the investigation being reported. | | 4 |
| *Objectives* | 3 | State specific objectives, including any pre-specified hypotheses | ***State if the study is the first report of a genetic association, a replication effort, or both.*** | 4 |
| **Methods** | | | | |
| *Study design* | 4 | Present key elements of study design early in the paper. | | Fig 1 |
| *Setting* | 5 | Describe the setting, locations and relevant dates, including periods of recruitment, exposure, follow-up and data collection. | | 4 |
| *Participants* | 6 | (a) **Cohort study –** Give the eligibility criteria, and the sources and methods of selection of participants. Describe methods of follow-up.<br><br>**Case–control study –** Give the eligibility criteria, and the sources and methods of case ascertainment and control selection. Give the rationale for the choice of cases and controls.<br><br>**Cross-sectional study –** Give the eligibility criteria, and the sources and methods of selection of participants. | ***Give information on the criteria and methods for selection of subsets of participants from a larger study, when relevant.*** | 5 |
| | | **(b) Cohort study** – For matched studies, give matching criteria and number of exposed and unexposed.<br><br>**Case–control study –** For matched studies, give matching criteria and the number of controls per case. | | |
| *Variables* | 7 | (a) Clearly define all outcomes, exposures, predictors, potential confounders, and effect modifiers. Give diagnostic criteria, if applicable. | ***(b) Clearly define genetic exposures (genetic variants) using a widely –used nomenclature system. Identify variables likely to be associated with population stratification (confounding by ethnic origin).*** | (a)5, Table S1<br>(b) 6,7 |

| | | | | |
|---|---|---|---|---|
| *Data sources measurement* | 8* | (a) For each variable of interest, give sources of data and details of methods of assessment (measurement). Describe comparability of assessment methods if there is more than one group. | ***(b) Describe laboratory methods, including source and storage of DNA, genotyping methods and platforms (including the allele calling algorithm used, and its version), error rates and call rates. State the laboratory /centre where genotyping was done. Describe comparability of laboratory methods if there is more than one group. Specify whether genotypes were assigned using all of the data from the study simultaneously or in smaller batches.*** | 5<br>6 |
| *Bias* | 9 | (a) Describe any efforts to address potential sources of bias. | ***(b) For quantitative outcome variables, specify if any investigation of potential bias resulting from pharmacotherapy was undertaken. If relevant, describe the nature and magnitude of the potential bias, and explain what approach was used to deal with this.*** | 5 |
| *Study size* | 10 | Explain how the study size was arrived at. | | 7 |
| *Quantitative variables* | 11 | Explain how quantitative variables were handled in the analyses. If applicable, describe which groupings were chosen, and why. | ***If applicable, describe how effects of treatment were dealt with.*** | 7 |
| *Statistical methods* | 12 | (a) Describe all statistical methods, including those used to control for confounding. | ***State software version used and options (or settings) chosen.*** | 7,8 |
| | | (b) Describe any methods used to examine subgroups and interactions. | | Table S1, 8 |
| | | (c) Explain how missing data were addressed. | | |
| | | (d) **Cohort study –** If applicable, explain how loss to follow-up was addressed.<br><br>**Case–control study –** If applicable, explain how matching of cases and controls was addressed.<br><br>**Cross-sectional study –** If applicable, describe analytical methods taking account of sampling strategy. | | |
| | | (e) Describe any sensitivity analyses. | | |

| | | | | |
|---|---|---|---|---|
| | | | ***(f) State whether Hardy-Weinberg equilibrium was considered and, if so, how.*** | 7, 8 |
| | | | ***(g) Describe any methods used for inferring genotypes or haplotypes.*** | 7 |
| | | | ***(h) Describe any methods used to assess or address population stratification.*** | 7 |
| | | | ***(i) Describe any methods used to address multiple comparisons or to control risk of false positive findings.*** | 7,8 |
| | | | ***(j) Describe any methods used to address and correct for relatedness among subjects.*** | 6 |
| **Results** | | | | |
| *Participants* | 13* | (a) Report the numbers of individuals at each stage of the study – e.g. numbers potentially eligible, examined for eligibility, confirmed eligible, included in the study, completing follow-up and analysed. | ***Report numbers of individuals in whom genotyping was attempted and numbers of individuals in whom genotyping was successful.*** | 6-7 |
| | | (b) Give reasons for non-participation at each stage. | | |
| | | (c) Consider use of a flow diagram. | | |
| *Descriptive data* | 14* | (a) Give characteristics of study participants (e.g. demographic, clinical, social) and information on exposures and potential confounders. | ***Consider giving information by genotype.*** | 10 |
| | | (b) Indicate the number of participants with missing data for each variable of interest. | | |
| | | (c) **Cohort study –** Summarize follow-up time, e.g. average and total amount. | | |
| *Outcome data* | 15* | **Cohort study –** Report numbers of outcome events or summary measures over time. | ***Report outcomes (phenotypes) for each genotype category over time*** | Fig 2 |
| | | **Case–control study –** Report numbers in each exposure category, or summary measures of exposure. | ***Report numbers in each genotype category*** | |
| | | **Cross-sectional study –** Report numbers of outcome events or summary measures. | ***Report outcomes (phenotypes) for each genotype category*** | |
| *Main results* | 16 | (a) Give unadjusted estimates and, if applicable, confounder-adjusted estimates and their precision (e.g. 95% confidence intervals). Make clear which confounders were adjusted for and why they were included. | | 7 |
| | | (b) Report category boundaries when continuous variables were categorized. | | |

| | | | |
|---|---|---|---|
| | | (c) If relevant, consider translating estimates of relative risk into absolute risk for a meaningful time period. | |
| | | ***(d) Report results of any adjustments for multiple comparisons.*** | Bonferroni thresholds given |
| *Other analyses* | 17 | (a) Report other analyses done – e.g. analyses of subgroups and interactions, and sensitivity analyses. | 10 Fig 2 B,C,E,F |
| | | ***(b) If numerous genetic exposures (genetic variants) were examined, summarize results from all analyses undertaken.*** | Fig 3, tables 1-3 |
| | | ***(c) If detailed results are available elsewhere, state how they can be accessed.*** | |
| **Discussion** | | | |
| *Key results* | 18 | Summarize key results with reference to study objectives. | 14 |
| *Limitations* | 19 | Discuss limitations of the study, taking into account sources of potential bias or imprecision. Discuss both direction and magnitude of any potential bias. | 17-18 |
| *Interpretation* | 20 | Give a cautious overall interpretation of results considering objectives, limitations, multiplicity of analyses, results from similar studies, and other relevant evidence. | 18 |
| *Generalizability* | 21 | Discuss the generalizability (external validity) of the study results. | 17-18 |
| **Other information** | | | |
| *Funding* | 22 | Give the source of funding and the role of the funders for the present study and, if applicable, for the original study on which the present article is based. | 19 |

STROBE: STtrengthening the Reporting of Observational Studies in Epidemiology

*Give information separately for cases and controls in case–control studies and, if applicable, for exposed and unexposed groups in cohort and cross-sectional studies.
